# Effective interventions to support recovery of people with psychosis and their families across socio-ecological levels in low-income and middle-income countries: a systematic review

**DOI:** 10.1101/2025.10.20.25338375

**Authors:** Sarah Barber, Lauren McPhail, Siqi Xue, Rachel Greenley, Cai Jia, Esubalew Assefa, Wubalem Fekadu, Awoke Mihretu, Hannah Weir, Roxanne C Keynejad, Emily West, Sudipto Chatterjee, Susan Cleary, Bonginkosi Chiliza, Julian Eaton, Charlene Sunkel, Craig Morgan, Ashok Malla, Charlotte Hanlon

**Author notes:** Joint senior authors (Ashok Malla, Charlotte Hanlon).

## Abstract

**Summary:** *Background:* The aim of this systematic review was to synthesise evidence on the effectiveness and cost-effectiveness of interventions to support the recovery of people living with psychosis and their families in low-income and middle-income countries (LMICs).

*Methods:* We searched nine databases for articles published from January 2001 to January 2024 without language restrictions. Studies were eligible if they enrolled people living with psychosis or family members, and tested a psychoeducational, psychological, social, economic or service intervention or delivery or implementation strategy aimed at improving outcomes of people with psychosis. Eligible studies were required to compare outcomes with an alternative condition, using any prospective evaluation study design in a LMIC setting. We extracted summary data from published papers and appraised risk of bias using the Effective Public Health Practice Project tool. We prioritised the reporting of recovery-orientated outcomes including social inclusion, personal recovery, reduced stigma and discrimination and human rights protections. We conceptualised the person living with psychosis in their context (individual, family, organisation and community) based on the socio-ecological model of disability and highlighted studies intervening and measuring outcomes across multiple socio-ecological levels. Protocol registration: PROSPERO (CRD42022330298).

*Findings:* A total of 310 individual studies including data from 34,435 participants in 37 countries were included. Aggregate data from a further five meta-analyses, comprising data from 130 individual studies were also included. The majority of studies (77%) were conducted in upper middle-income countries. There was a dominance of studies evaluating impacts of interventions on individual-level mental health and functioning and a paucity of studies measuring the recovery-orientated outcomes prioritised by people living with psychosis. There were modest effects for comprehensive interventions involving family, psychosocial rehabilitation and care close to home provided by trained specialists however their scalability in resource-limited settings is unclear. Over half the studies were considered to have a high risk of bias.

*Interpretation:* There is a need for studies that evaluate scalable interventions supporting recovery with comprehensive and contextualised outcome measures and for greater investment in strengthening capacity to conduct rigorous psychosis research across LMICs.

*Funding:* None.

**Research in context:** *Evidence before this study:* Recent World Health Organization (WHO) guidance on human rights-based, recovery-orientated community mental health care featured markedly few case studies of good practice for people with psychosis in low-income and middle-income countries (LMICs). Systematic reviews of interventions for psychosis in LMICs have been narrow in focus and reporting outcomes, and limited to English language publications.

*Added value of this study:* This systematic review is the most comprehensive synthesis to date of psychosis interventions in LMICs. Inclusion is not restricted by publication language. We highlight studies reporting recovery-oriented outcomes prioritised by people living with psychosis and impacts of interventions across levels of the socio-ecological model of disability. While being particularly relevant to LMICs, our findings also contribute a useful perspective for high income settings.

*Implications of all the available evidence:* Most interventions were targeted at the individual and focused on mental health and functioning outcomes, with few evaluations of impact on social inclusion and other valued outcomes. There is some evidence in support of specialist-delivered comprehensive interventions involving family, psychosocial rehabilitation and care close to home, but effect sizes were small-to-modest, and many intervention types and delivery agents have not been adequately tested, especially in LICs and rural settings. There is a clear need to develop comprehensive and contextualised measures for recovery-orientated outcomes and to invest in strengthening capacity to conduct rigorous research on interventions for psychosis in LMICs.

## Introduction

An estimated 3-3·5% of the global adult population experiences psychosis in their lifetime. Psychotic disorders (‘psychoses’) include schizophrenia and affective conditions in which psychotic episodes may be recurrent or enduring, resulting in distress and/or functional impairment.^1^ Early multi-country studies indicated better clinical outcomes for people with psychosis in some low-income and middle-income countries (LMICs) than in high-income countries (HICs), purportedly due to living in more supportive sociocultural environments.^2^ These early findings were largely not reproducible, with evidence from diverse LMICs showing that psychoses can have profound adverse impacts on individuals, including excess mortality, human rights abuses, impoverishment and social exclusion, alongside distress from the experience of psychosis.^3-6^ Due to poor state support, access to formal mental health services and social welfare programmes in LMICs is often scarce, and families (including unrelated informal caregivers) play crucial roles in providing care. This means that families often share the adverse impacts of psychosis, which have been shown to extend across generations.^7^

In recent years, in largely western settings, there have been calls to move interventions and services for people with psychoses from symptom-based to those supporting recovery-orientated goals.^8^ Recovery-orientated interventions aim to support people on their unique recovery journeys, helping to pursue goals that matter to them personally.^8^ The widely adopted ‘CHIME’ recovery model, developed through a synthesis of published descriptions and models of personal recovery, defines five key elements of personal recovery: connectedness, hope and optimism, identity, meaning and purpose, and empowerment.^8^

However, understandings of what recovery from psychosis means varies across sociocultural contexts.^9^ The ‘CHIME’ model was developed from studies conducted predominantly in the USA, UK, Australia and Canada^8^. Research has shown that the extent of societal interdependence, religiosity and spirituality, economic precarity and access to care and treatment are important influences on how individuals and their families frame recovery processes.^10^ Social inclusion, or the extent to which people with psychosis and their families are able to meaningfully participate in aspects of community life as equal citizens, may resonate more than personal recovery concepts of self-actualisation or individual empowerment in more collectivistic settings.^11^ Indeed, studies across diverse settings support the prioritisation of social inclusion as a valued outcome.^12^ However, key components of social inclusion require clarification,^13^ and vary between settings.^14^

Efforts to enhance the participation of people living with psychosis presume the existence of a desirable, identifiable social space within which the individual expects to be included.^14^ Yet social environments can be hostile due to widespread stigma and discrimination against people living with psychosis and their families.^15^ Although the family home can be a place of shelter and protection for people living with psychosis, family care can at times be excluding, discriminatory or coercive.^14^ Whether interventions succeed in supporting recovery should therefore be evaluated in terms of their impacts not just at the individual level, but also at levels of their family environment, the organisations and services with which they interact, and their communities.^16^ This contextual and socially orientated lens has not yet been applied to the existing evidence on psychosis interventions in LMICs.

A recent scoping review of literature published in English from 2012-2022 described multi-sectoral and inter-sectoral approaches to enabling recovery from psychosis in LMICs.^17^ The authors identified 36 records and themes of collaboration between health care and community support systems, supported housing and supportive community spaces for recovery. However, the authors did not consider definitions of recovery or synthesise evidence of effectiveness. Recent World Health Organization (WHO) *Guidance and technical packages on community mental health services: promoting person-centred and rights-based approaches* featured markedly few case studies of good practice for people with psychosis in LMICs.^18^

The aim of this systematic review was to synthesise evidence on the effectiveness and cost-effectiveness of interventions to support the recovery of people with psychosis and their families across levels of the socio-ecological model of disability in LMICs. We prioritised outcomes relating to personal recovery and social inclusion, and highlighted studies measuring reductions in stigma and discrimination and human rights violations, as these are universal barriers to achieving the outcomes that matter most to people living with psychosis.

## Methods

We followed the updated PRISMA reporting guideline.^19^ The protocol was registered with PROSPERO on 18 May 2022 with minor amendments published on 1 Nov 2022 (CRD42022330298). Further minor deviations from the protocol are detailed in supplementary file 1.

### Information sources and search strategy

We searched nine databases: Embase, Global Health, Global Index Medicus, MEDLINE, PubMed, PsycINFO, Scopus, Web of Science, and Tufts Medical Center Cost-Effectiveness Analysis (CEA) Registry, for publications dating from January 2001, without language restrictions. Searches were run on 5^th^ June 2022 and updated on 10^th^ January 2024.

Database-specific searches included keywords and MESH terms for psychosis combined with an extensive list of search terms for interventions. We combined these using Boolean operators with terms for LMICs and eligible study designs (see supplementary file 2). Studies were identified through electronic database and hand searches of references in relevant systematic reviews.

### Eligibility criteria

Studies were eligible for inclusion if they were conducted in countries categorised by the World Bank as an LMIC in the year of publication, and fulfilled the following criteria:

### Participants

People with psychosis (aged 14+ years) or individuals (family/non-family) involved in the unpaid care of people living with psychosis (‘family members’), where a diagnosis of non-affective or affective psychosis was made using standardised diagnostic criteria or clinical assessment. Studies that included people with diverse mental health conditions were eligible if either: more than 50% of the study sample had a primary diagnosis of psychosis, or outcomes for participants with psychosis were reported separately. Studies of people from LMICs who had migrated to a high-income country, or persons at risk or with prodromal or sub-clinical symptoms were not eligible.

### Intervention

The review team developed a comprehensive list of search terms for interventions.^20^ These included anti-stigma, cognitive, creative art therapies, family intervention, livelihood and employment, peer support, physical health, psychiatric care, psychoeducation, psychological therapy, rehabilitation and recovery, service-level, social and substance use interventions and traditional practices.

We excluded efficacy or effectiveness studies of any medicinal compounds (pharmaceutical, homeopathic, herbal) or other biomedical interventions (e.g. electroconvulsive therapy) but included interventions to support medication adherence (strategies combined with the administration of medication). All study participants were assumed to have been treated with antipsychotic medication where needed if not explicitly stated. Strong ethical justification would be required if an intervention was provided instead of, rather than as an adjunct to antipsychotic treatment.

### Context

We included studies conducted in any LMIC context, including community settings, prisons, residential settings, traditional or religious healing sites and healthcare settings. However, in the synthesis of findings, we prioritised reporting interventions that did not rely on a specialist inpatient platform for delivery in line with the WHO guidance on community-oriented services.

### Outcomes

Prioritised outcomes were: personal recovery (including measures of hope and empowerment), social inclusion, reduced stigma and discrimination, and/or human rights protections.

Beyond this, we extracted data on a range of outcomes conceptualised as on a pathway to our prioritised outcomes, including outcomes related to domains of the WHO community-based rehabilitation framework^21^ (i.e. mental and physical health; social functioning and support; economic functioning and livelihoods) and access to services. Recognising the central role of family in LMIC contexts, we also extracted outcomes relating to the family unit (e.g., functioning, communication, burden) as well as individual family member outcomes (e.g. mental health, physical health). We also extracted individual, household and service or system level cost data in order to assess evidence of cost-effectiveness, including incremental cost effectiveness ratios (ICERs) (See supplementary file 3 for a full list of categories).

We mapped all of the above outcome domains to the socio-ecological model of disability,^16^ recognising that interventions can impact different socio-ecological levels simultaneously (Figure 1). Where included studies collected outcome data across multiple time points, we made a pragmatic decision to extract data from the first post-intervention timepoint available.

**Figure 1.**
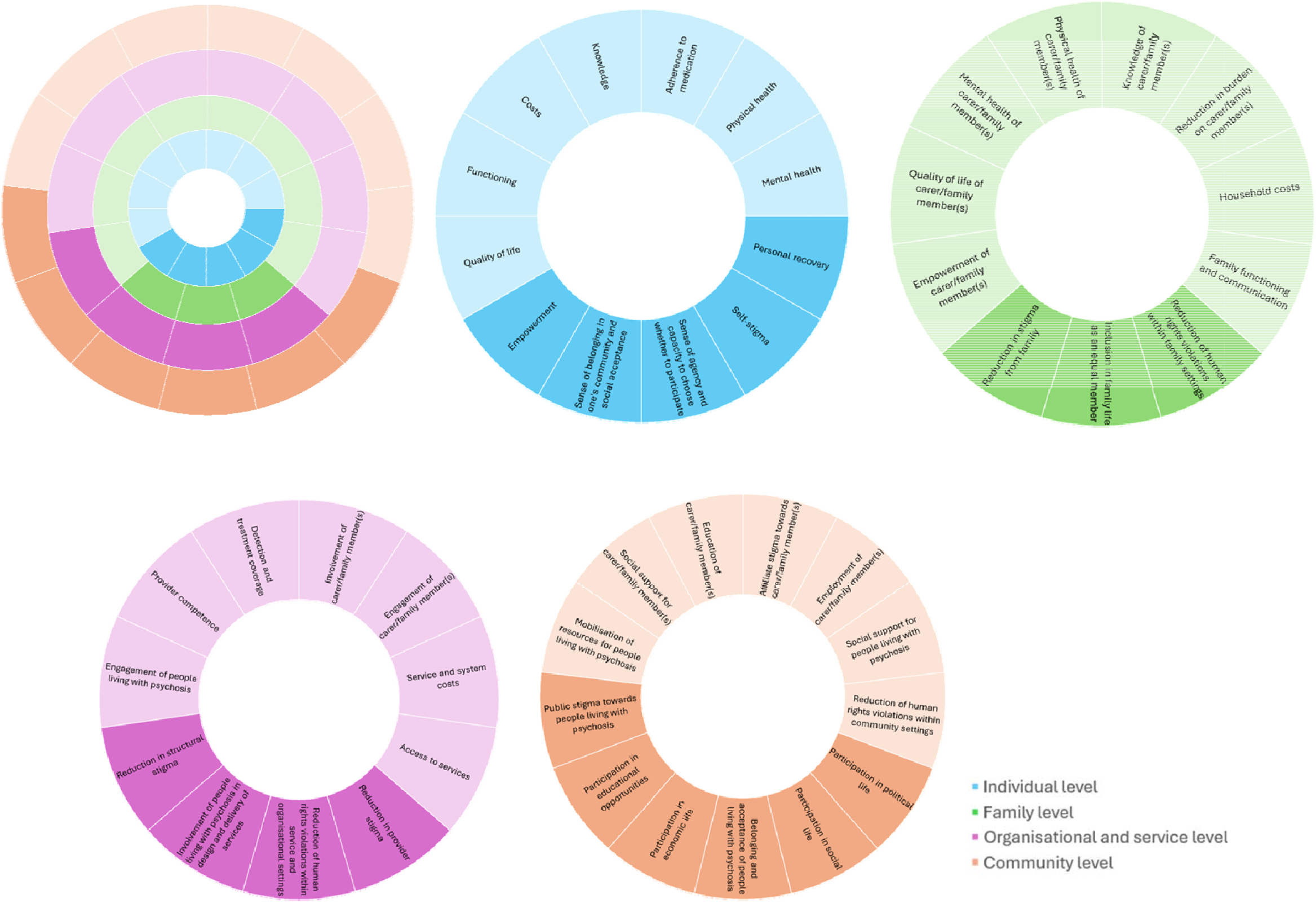
Outcomes mapped across levels of the socioecological model (priority outcomes highlighted)

### Study design

Eligible studies were peer-reviewed scientific publications. Eligible designs were any longitudinal intervention study or synthesis, including meta-analyses, randomised controlled trials (RCTs; individual and cluster), controlled clinical trials, two group pre-post comparisons, uncontrolled before-after and interrupted time series. Pilot studies and feasibility designs were included in order to meet the secondary objective of mapping the intervention landscape, but not to contribute to the effectiveness assessment. Mixed-methods studies were eligible for inclusion if quantitative data could be extracted. Full and partial economic evaluations were included provided that the study design met all inclusion criteria.

We excluded experimental studies not clearly linked to real world impacts (e.g. examining an isolated cognitive process). Qualitative evaluations, conference abstracts, posters and student theses were excluded.

### Selection process

Titles and abstracts were initially double screened independently by researchers (AM, CJ, EW, HW, RCK, RG, SB, SX, WF) working in pairs using the online platform Rayyan^22^. Disagreements were resolved through discussion amongst reviewers, including senior authors (CH, AKM). Once at least 500 records had been double-screened and >80% agreement was achieved between pairs, we proceeded to single screening, involving senior authors for any uncertainties.

Full texts were located, and non-English papers were translated using Google Translate, except for Chinese, Thai and Persian papers, which were translated by native speakers. Full texts were double screened for inclusion against the eligibility criteria by three authors (RCK, RG, SB) and discussed with a senior author if required.

### Data collection process and data items

Data from included studies were first extracted independently by one reviewer (AM, CJ, HW, LM, PM, RCK, RG, SB, SF, SX, WF) into a standardised and pre-piloted table (Supplementary file 4). Data from eligible individual studies identified in published Cochrane Reviews were extracted from summary tables due to time and resource constraints. All extracted data were double checked by two reviewers (SB, LM), either against the original paper or, in the case of non-English language papers, by seeking clarifications from the extractor.

For priority outcomes, we extracted effect sizes (where reported) or data that allows for a judgement of effect size, alongside p-values. For all other outcomes, we extracted and categorised outcome data based on the level of statistical significance of reported differences between comparison conditions, as follows: statistically significant (p<0·05) and in the expected direction for an effective intervention (classified as ‘SD’), not statistically significant (NS), or significantly different in an unexpected direction (SD-). We did not extract data from sub-scales, except for the Positive and Negative Symptoms Scale (PANSS), ^23^ where we delineated impacts on positive versus negative symptoms of psychosis, when data were available.

### Risk of bias assessment

9

We used the Effective Public Health Practice Project (EPHPP) tool for quantitative studies to evaluate the risk of bias of individual studies contributing to the evidence of effectiveness. ^24^Studies were assigned global ratings of ‘strong’ (low risk of bias), ‘moderate’ (moderate risk of bias) or ‘weak’ (high risk of bias), based on assessments of sub-domains. Consistent EPHPP ratings were confirmed through double rating of a subset of papers and discussion with senior authors (AKM, CH), supported by twice-monthly online meetings during the rating period. Studies with fewer than 30 participants at baseline without a hypothesis-driven sample size calculation were automatically assigned high risk of bias. We used the AMSTAR-2 tool to evaluate the risk of bias of included meta-analyses.^25^

### Effect measures

We did not calculate meta-analytic summaries of intervention effects due to the heterogeneity of outcome measures reported.

### Synthesis methods

We present aggregated summaries of descriptive data. We confined our synthesis of effectiveness findings to studies which were not pilot, feasibility or head-to-head designs and which were classified as being at low or moderate risk of bias. We present evidence of effectiveness for (1) prioritised outcomes and (2) studies reporting outcomes across multiple socio-ecological levels, limiting results in the main manuscript to studies outside of psychiatric inpatient settings.

## Results

Our search identified 43,333 records (Figure 2). After removing duplicates, we screened 28,040 titles and abstracts. A total of 579 records were included for full-text review, of which 324 records reporting on 315 studies were eligible for inclusion. We extracted data from 310 individual studies (of which 127 were randomised,183 were non-randomised, and 22 were pilot or feasibility designs) reporting data from 34,435 participants in 37 countries. We additionally extracted pooled results from five meta-analyses of RCTs of narrowly defined interventions implemented in China (n=4) and China/Tunisia (n=1), summarising a further 131 individual studies with over 6,358 participants.

**Figure 2.**
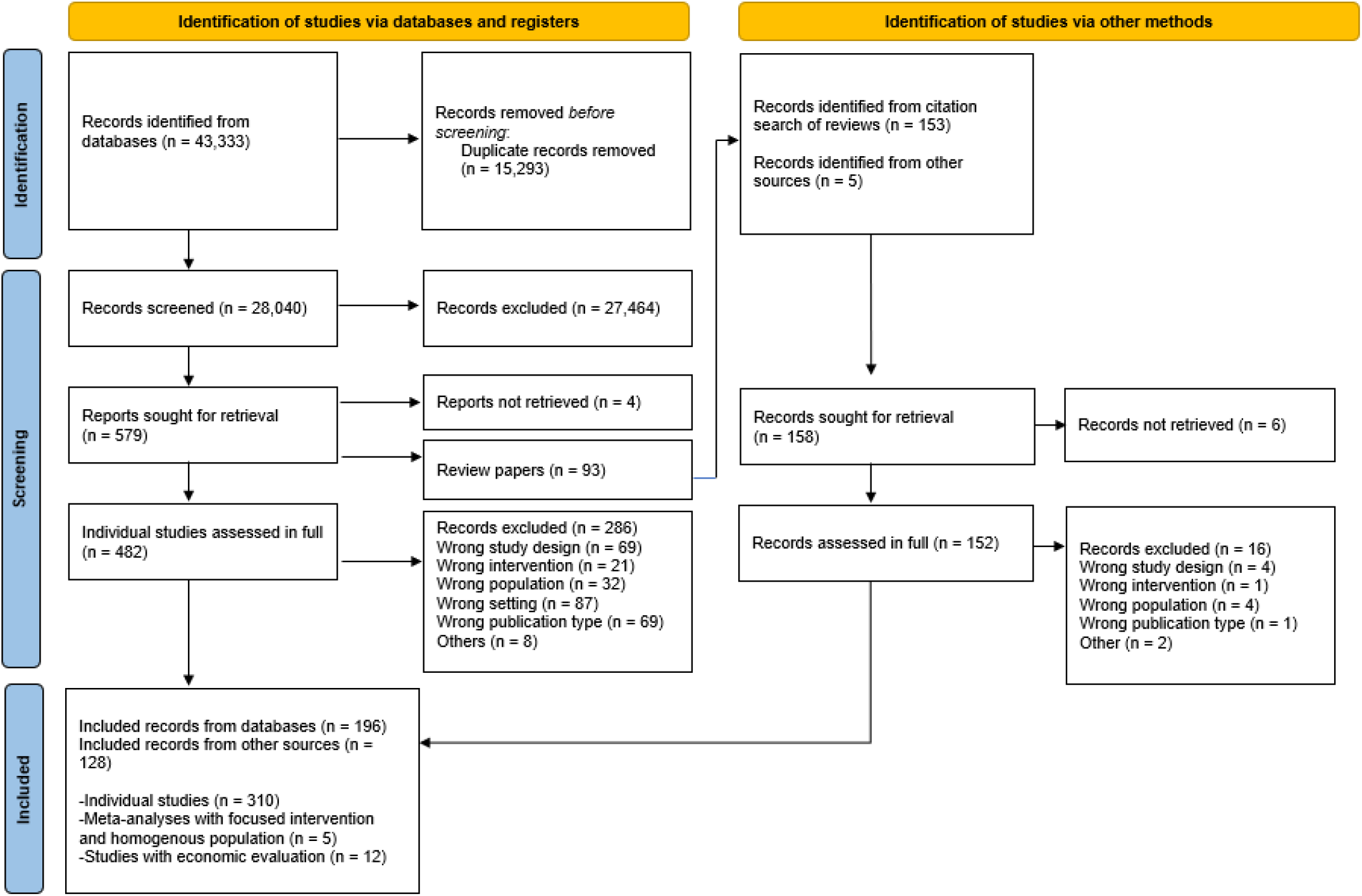
PRISMA Flow Diagram

Characteristics of included studies are summarised in Table 1. The majority of studies were from upper-middle-income countries, with just 3% (9/315) from low-income countries. The largest grouping by WHO region was the Western Pacific Region, accounting for 45% (141/315), of which 137 studies were conducted in China. Most studies were conducted in urban settings and relied on a hospital-based platform for intervention delivery. Only 8% (26/315) of studies explicitly reported being conducted in rural settings, and just 17% (55/315) of studies used a community platform.

**Table 1.** Characteristics of included studies (n=315)

| **Setting** | **N (%)** |
| --- | --- |
| *Country income classification* |  |
| Low income | 9 (3) |
| Lower middle income | 62 (20) |
| Upper middle income | 243 (77) |
| Mixed income levels | 1 (0) |
| *World Health Organization Region* |  |
| African Region | 18 (6) |
| Eastern Mediterranean Region | 40 (13) |
| European Region | 25 (8) |
| Region of the Americas | 17 (5) |
| South-East Asia Region | 73 (23) |
| Western Pacific Region | 141 (45) |
| Mixed regions | 1 (0) |
| *Within country setting* |  |
| Urban | 212 (67) |
| Rural | 26 (8) |
| Mixed | 22 (7) |
| Not specified | 55 (17) |
| **Study design** |  |
| Meta-analysis of randomised control trials | 5 (2) |
| Randomised control trial (individual) | 112 (36) |
| Randomised control trial (cluster) | 9 (3) |
| Controlled clinical trial | 97 (31) |
| Two group pre-post study | 22 (7) |
| Uncontrolled before-after study | 47 (15) |
| Interrupted time series study | 1 (0) |
| Pilot or feasibility study | 22 (7) |
| **Risk of bias\*** |  |
| Low risk | 53 (18) |
| Moderate risk | 81 (28) |
| High risk | 159 (54) |
| **Intervention components (some interventions comprised multiple components)** |  |
| Psychoeducation ((people living with psychosis) | 116 (37) |
| Psychoeducation (family/caregiver) | 102 (33) |
| Social or rehabilitation | 88 (28) |
| Psychological therapy (people living with psychosis) | 73 (23) |
| Psychological therapy (family/caregiver) | 53 (17) |
| Psychiatric care | 31 (10) |
| Cognitive | 22 (7) |
| Physical health | 17 (5) |
| Creative arts therapies | 13 (4) |
| Peer or mutual support (family/caregiver) | 12 (4) |
| Peer or mutual support (people living with psychosis) | 10 (3) |
| Economic or educational | 11 (3) |
| Awareness or anti-stigma | 9 (3) |
| Traditional practices | 5 (2) |
| Service-level | 54 (17) |
| System-level | 4 (1) |
| **Platform for intervention** |  |
| Community | 55 (17) |
| Social care/community residential | 6 (2) |
| Primary health care/community health centre | 11 (3) |
| Community mental health service | 10 (3) |
| General hospital out-patient/inpatient/not-specified | 6 (2) |
| Psychiatric hospital out-patient | 68 (22) |
| Psychiatric hospital in-patient | 70 (22) |
| Psychiatric hospital in-patient and out-patient | 3 (1) |
| Psychiatric hospital not specified | 10 (3) |
| Hospital (not-specified) out-patient/in-patient/not-specified | 34 (11) |
| Mixed hospital/community (e.g. home visits from a hospital) | 16 (5) |
| Prison | 2 (1) |
| Other | 3 (1) |
| Not specified | 21 (7) |
| **Delivered by** |  |
| *Any mental health professional\*\** | 190 (60) |
| Psychiatrist/ psychologist | 118 (37) |
| Psychiatric nurse/ clinical officer/ not specified | 80 (25) |
| Allied professional (specialised) | 59 (19) |
| *Any general health or allied professional\*\** | 32 (10) |
| Physician | 10 (3) |
| Non-physician (facility based) | 11 (3) |
| Allied professional (generalist) | 13 (4) |
| *Any community health worker\*\** | 18 (6) |
| Community mental health worker | 3 (0) |
| Community health worker | 15 (5) |
| Lay worker or volunteer | 19 (6) |
| Automated or digital | 13 (4) |
| Peer | 8 (3) |
| Family member or informal caregiver | 8 (3) |
| Traditional or faith healer | 4 (1) |
| Police or justice worker | 1 (0) |
| Researcher (not specified) | 23 (7) |
| Not specified | 70 (22) |

\*Excludes pilot/feasibility studies (n=22), \*\*Can be multiple

Most studies (82%; 258/315) enrolled only people living with psychosis, 8% (26/315) enrolled only family members and 10% (31/315) enrolled both. Total baseline sample sizes of individual studies (excluding pilot or feasibility designs) ranged from 10 to 1268 (mean=115·3, median=95). The proportion of participants with psychosis who were female was 40% (gender balance was reported in 227/289 studies) and mean age was 36·1 years (reported in 210 of 289 studies). From available data on duration of illness in 149 studies, 12% (18/149) of studies enrolled participants with predominantly early (0-2 years duration) psychosis, whilst 42% (63/149) enrolled participants with predominantly longer term (10 years or more duration) psychosis.

The largest number of single component interventions comprised psychoeducation for people with psychosis (20%; 31/158), followed by social or rehabilitation interventions (13%; 21/158) and psychological therapies for people living with psychosis (12%; 19/158) or family (12%, 19/158) and psychoeducation for family (11%; 18/158). In the 50% (157/315) of studies with multiple components, the aforementioned intervention types were also the most frequently evaluated. Other intervention types studied either alone or in combination included psychiatric care, peer or mutual support, economic or livelihood interventions, anti-stigma, creative arts, physical health and traditional practices. Specialist mental health workers were involved in the delivery of the intervention in 60% (190/315) of studies. Only 10% (32/315) of interventions involved general health workers, such as generalist nurses or physicians, 6% (18/315) involved non-specialist community health workers, 3% (8/315) involved peers and 2% involved family members (7/315).

The most commonly measured outcome domains across all studies were at the individual level: mental health symptoms (62%; 197/315), functioning (38%; 118/315), adherence to treatment (20%; 62/315) and quality of life (13%; 41/315), followed by family burden (11%; 33/315) and family functioning and communication (7%; 21/315) at the family level and engagement (or satisfaction) of family (6%; 20/315) with services, at the organisational level. The remaining outcome domains were each measured in less than 5% of studies. The frequency of outcome reporting is shown in Figure 3 and described in detail in Supplementary file 3.

**Figure 3.**
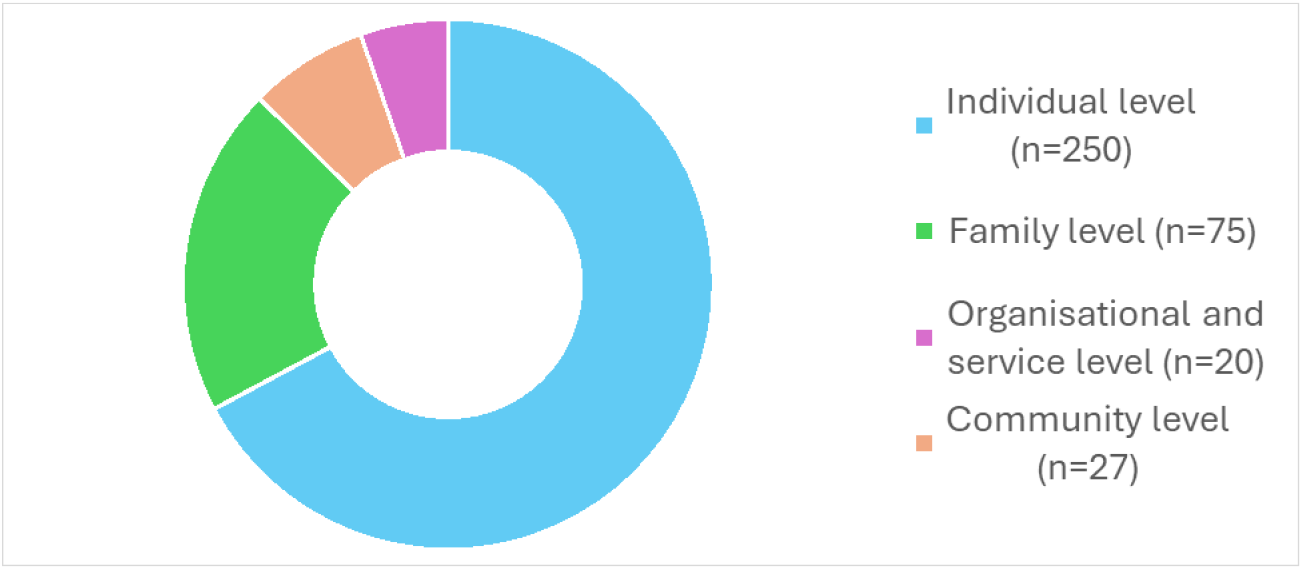
Frequency of reporting at least one outcome per domain across studies (n=315)

The results of 16 studies reporting prioritised outcomes with moderate-low risk of bias are summarised in Table 2 (See supplementary file 5 for findings from all studies). We found some evidence that multi-component interventions for people living with psychosis and/or family members delivering psychoeducation, family intervention and additional components (either skills training and cognitive behavioural therapy, or crisis intervention), resulted in modest benefits for measures of social inclusion, specifically a 10% increase access to education or employment^26^ and 10% reduction in inability to work ^27^ in two studies in China. Community health worker-led case management in Thailand resulted in a small, non-significant increase in percentage employed to 3%.^28^

**Table 2.** Studies measuring prioritised outcomes with moderate-low risk of bias (excluding inpatient (psychiatric/not-specified) settings)

| Author | Year | Country | Design | Baseline total | Intervention | Platform | Primary outcome | Social inclusion | Personal recovery | Reduction in stigma and discrimination | Human rights protections | Quantitative data (p-value, effect size or other) |
| --- | --- | --- | --- | --- | --- | --- | --- | --- | --- | --- | --- | --- |
| Guo et al<sup>26</sup> | 2010 | China | RCT (individual) | 1268 | Manualised group psychosocial intervention once monthly for one year, comprising psychoeducation, family intervention, skills training and cognitive-behavioural therapy delivered by mental health professionals | Specialist hospital out-patient | Rate of or time to treatment discontinuation or change | SD | - | - | - | Access to education or employment significantly higher in intervention vs control group at one-year follow up, 30·1% (intervention) vs 22·2% (control) ($\chi^2=10·094$, $p=0·001$).<br>Effect size: not reported. |
| Ran et al<sup>27</sup> | 2015 | China | RCT (cluster) | 238 | Family psychoeducation once a month for nine months, family workshops every three months and crisis intervention delivered by trained psychiatrists assisted by 'village doctors' | Community | Not clearly stated | SD | - | - | - | 'Work ability' significantly different between groups at 14-year follow up e.g. 'Unable to work' = 21·1% (intervention) vs 30·6% (depot medication only) vs 42·3% (TAU) ($\chi^2=11·4$, $p=0·022$), pairwise comparisons not reported<br>Effect size: not reported. |
| Jirapramukpitak et al<sup>28</sup> | 2022 | Thailand | Two group pre-post | 549 | Community health worker-led assertive case management with support from specialist hospital staff (psychiatrist, clinical psychologist, nurse) | Community | Employment status and service use | NS | - | - | - | No significant difference between groups in employment at one-year follow up ($p=0·686$)<br>Effect size: risk difference = 3% (95% CI -11% to 17%) |
| Muhic et al<sup>29</sup> | 2022 | Bosnia and Herzegovina | RCT (individual) | 72 | Multi-family group meetings between people living with psychosis and one or two selected family members or friends, facilitated by mental health professionals, meeting once a month for six months | Specialist hospital out-patient | Quality of life | NS | - | - | - | No significant difference between groups in objective social outcome (SIX) score at 6-month follow up, mean score (SD) = 4·5 (1·2) (intervention) vs 4·1 (1·3) (control) ($p=0·105$)<br>Effect size: Proportional OR = 0·976; Cohen's $d$ = 0·31 |
| Sikira et al<sup>30</sup> | 2021 | Bosnia and Herzegovina | RCT (individual) | 65 | Volunteer befriending programme | Community | Quality of life | NS | - | - | - | No significant difference in objective social outcome (SIX) score at 6-month follow up, mean score (SD) 4.2 (1.2) (intervention) vs 4.2 (1.5) (control) (p=0.072)<br>Effect size: proportional OR =1.48 (95% CI -0.13 to 3.08) |
| Asher et al<sup>31</sup> | 2022 | Ethiopia | RCT (cluster) | 332 | Community-based rehabilitation intervention lasting nine months and involving home visits by community health workers | Community | Disability | SD- | - | NS | NS | (1) Unemployment significantly higher in intervention vs control group at 12-month follow up, 70% (intervention) vs 62% (control) (p=0.034)<br>Effect size: adjusted OR 3.80 (95% CI 1.10 to 13.07)<br>(2) No significant difference in any experience of discrimination in the last six months between groups at 12-month follow up, 63% (intervention) vs 51% (control) (p=0.089)<br>Effect size: adjusted OR 2.26 (95% CI 0.88 to 5.81)<br>(3) No significant difference in physical restraint in the last 6 months at 12-month follow up, 7% (intervention) vs 7% (control) (p=0.35)<br>Effect size: adjusted OR 2.72 (95% CI 0.34 to 21.83) |
| Zhao et al<sup>32</sup> | 2013 | China | RCT (individual) | 105 | Monthly 40–60 minute online consultations with a mental health professional or web-based psychoeducation for people living with psychosis and caregivers | Community |  | - | - | SD | - | Statistically significant difference in stigma scale score between intervention and control group, mean score (SD) 35·77 (8·89) (intervention) vs 40·35 (12·24) (control) (p<0·05)<br>Effect size: not reported |
| El-Nahas et al<sup>33</sup> | 2018 | Egypt | RCT (individual) | 60 | Adapted manualised family intervention over six months (total 14 sessions) delivered by the researcher, including psychoeducation, communication enhancement, problem-solving skills | Specialist hospital out-patient | Psychosis symptom severity | - | - | SD | - | Significantly higher total Community Attitudes to the Mentally Ill (CAMI) scale score (indicating more positive attitudes) in intervention group compared to control at 6-months, mean score (SD) 65·4 (6·6) (intervention) vs 54·0 (6·1) (control) (p<0·001)<br>Effect size: not reported |
| Hasan et al<sup>34</sup> | 2017 | Jordan | RCT (individual) | 112 | Empowerment component of a comprehensive group rehabilitation intervention for people living with psychosis, delivered by the researcher in weekly sessions for six weeks | Specialist hospital out-patient | Helplessness | - | SD | - | - | Significant improvement in Recovery Assessment Scale (RAS) score post-treatment in empowerment group vs TAU at 6-weeks, mean score (SD) 55.45 (7.67) at baseline and 88.56 (9.65) at end-point (intervention) vs 56.37 (6.45) at baseline and 56.01 (6.34) at end-point (p<0.001)<br>Effect size: not reported |
| Tao et al<sup>35</sup> | 2021 | China | Meta-analysis | MA (1749 across included studies) | Mindfulness meditation (n=20 studies) | Mixed | N/A | - | SD | SD | - | (1) Pooled effect size (Hedge's *g*) for self-efficacy = 0.646 (95% CI 0.378 to 0.914), p<0.001 and self-esteem = 0.599 (95% CI 0.332 to 0.867), p<0.001<br>(2) Pooled effect size (Hedge's *g*) for perceived stigma = 0.813 (95% CI 0.282 to 1.067), p=0.001 |
| Gureje et al<sup>36</sup> | 2020 | Ghana, Nigeria | RCT (cluster) | 307 | Collaborative care model involving traditional faith healers and primary care health workers | Primary health care/ community health centre | Psychosis symptom severity | - | - | SD | NS* | (1) Significant effect of intervention on Internalised Stigma of Mental Illness (ISMI) scale score at 6-months follow-up, mean score (SD) 2.0 (0.6) (intervention) vs 2.1 (0.6) (control) (p=0.007)<br>Effect size: adjusted mean difference -0.2 (95% CI -0.3 to 0.0)<br>(2) No significant difference in change in proportion reporting harmful treatment practices between intervention and enhanced care as usual at 6-months, -48% (intervention) vs -33% (control) (p=0.071)<br>Effect size: difference in change in proportion between groups -15% (95% CI -32% to 1%) |
| Attux et al<sup>37</sup> | 2013 | Brazil | RCT (individual) | 160 | Lifestyle intervention involving physical health and nutrition, delivered by mental health professionals once weekly (60 minute) for 12 weeks | Specialist hospital out-patient | Weight and body mass index changes | - | NS | - | - | No significant difference in Rosenberg self-esteem (RSE) scale score between intervention and control groups at 3-months, mean score (SD) 12·1 (4·3) at baseline and 11·8 (4·8) at 3-months (intervention) vs 12·3 (5·0) at baseline and 12·2 (5·0) at 3-months (p=0·811)<br>Effect size: not reported |
| Fan et al<sup>38</sup> | 2022 | China | Uncontrolled before-after study | 167 | Community-based peer support | Community | Not clearly stated | - | NS | - | - | No significant difference in Chinese version of Rosenberg self-esteem (RSE) scale score at one-year follow-up, mean score (SD) 28·77 (5·29) (before) vs 29·96 (5·19) (one-year) (p=0·108)<br>Effect size: partial eta squared = 0·035 |
| Li et al<sup>39</sup> | 2018 | China | RCT (cluster) | 384 | A comprehensive intervention delivered by mental health professionals comprising psychoeducation, social skills training and cognitive behavioural therapy | Specialist hospital out-patient | Not clearly stated | - | NS | NS | - | (1) No significant effect of intervention on Self-esteem Scale (SES) score at 9-month follow up, mean score (SD) 22.95 (3.92) at baseline and 22.89 (3.61) at 9-months (intervention) vs 23.34 (3.88) at baseline and 23.23 (2.77) at 9-months (control) (p=0.805)<br>Effect size: regression co-efficient (*b*) =-0.14 (95% CI -1.25 to 0.97)<br>(2) No significant effect of intervention on Internalised Stigma of Mental Illness (ISMI) scale score at 9-month follow up, mean score (SD) 2.30 (0.38) at baseline and 2.24 (0.37) at 9-months (intervention) vs 2.30 (0.40) at baseline and 2.30 (0.39) at 9-months (control) (p=0.440)<br>Effect size: regression co-efficient (*b*) =-0.04 (95% CI -0.14 to 0.06) |
| Chatterjee et al<sup>40</sup> | 2014 | India | RCT (individual) | 282 | Community-based rehabilitation intervention lasting 12 months and involving home visits by community health workers | Community | Change in symptoms and disability | - | - | NS | - | No significant effect of intervention on proportion with any negative discrimination, 30% (intervention) vs 30% (TAU) or anticipated discrimination, 40% (intervention) vs 33% (TAU) at 12-months (p values not reported, see CI below)<br>Effect size: any negative discrimination OR 1.02 (95% CI 0.54 to 1.92); anticipated discrimination OR 1.31 (95% CI 0.66 to 2.60) |
| Ofori-Atta et al<sup>41</sup> | 2018 | Ghana | RCT (individual) | 143 | Psychiatric care combined with prayer camp treatment | Community | Psychiatric symptom severity | - | - | - | NS | No significant difference in the number of days spent restrained in chains between groups at 6 weeks follow-up, mean days 10.37 (intervention) vs 11.38 (control) (p=0.38)<br>Effect size: Cohen's *d* - 0.17 |

CI = confidence interval; MA = meta-analysis; N/A = not applicable; NS = non-statistically significant difference; OR = odds ratio; RCT = randomised controlled trial; SD = standard deviation; SD = statistically significant difference; SD- = statistically significant difference (unexpected direction); TAU = treatment as usual.
\*Both showed reductions

A study of multi-family group support meetings in Bosnia and Herzegovina found no meaningful improvement in objective social outcome score, a proxy for social inclusion.^29^ In comparison, a study of volunteer befriending reported non-significant, modestly higher odds of better social outcome with the same measure, however with small sample size and wide uncertainty.^30^ A study of community-based rehabilitation in Ethiopia found an unfavourable association with unemployment (OR 3·80) in a fully adjusted model.^31^

A six-week empowerment intervention in Jordan^34^ was associated with a significant improvement in a measure of personal recovery, although no effect size was reported. A meta-analysis of mindfulness meditation studies in China reported moderate-to-large effects of the intervention on aggregated measures of self-efficacy and self-esteem, as well as perceived stigma.^35^ There was no evidence that a multi-component community intervention^41^ or community peer-support in China^38^ meaningfully impacted on self-esteem^38,39^ or self-stigma.^39^ In comparison, a collabative care model involving traditional faith healers and primary care workers in Ghana and Nigeria resulted in a modest improvement in self-stigma scores.^36^

One study of an adapted family intervention in Egypt showed a modest benefit to caregiver attitudes towards people living with mental illness between groups after six months^33^, however did not report outcomes from the perspective of the person living with psychosis. Experience of discrimination was measured in two studies of community-based rehabilitation in Ethiopia^42^ and India^40^ with neither showing beneficial effects.

A study of collaborative care involving traditional faith healers and primary care health workers in Ghana and Nigeria demonstrated a large reduction in proportion reporting harmful treatment practices of almost 50%, although this was not significantly different to enhanced treatment as usual (-33%).^36^ Neither community based rehabilitation in Ethiopia^31^ nor collaborative care within prayer camps in Ghana^41^ impacted beneficially on physical restraint.

Overall, 55% (11/20) of studies with moderate-low risk of bias measuring prioritised outcomes across all settings reported statistically significant differences between comparison conditions favouring the intervention. Most studies had only small-to-modest effects on these valued outcomes.

In contrast, for individual mental health, functioning, quality of life and adherence to medication outcomes, 91% (91/100), 79% (53/67), 77% (20/26) and 81% (21/26) of studies found significant differences favouring the intervention respectively (Supplementary file 3).

Table 3 summarises the results of 30 studies reporting outcomes across multiple levels of the socio-ecological model with moderate-low risk of bias (see supplementary file 6 for all studies). Studies of interventions involving family, such as psychoeducation^27,32,34,46,47,50^, mutual support^44^ or behaviour change^51^ reported significant differences across multiple levels (e.g. individual functioning, family burden and social support), as did a number of rehabilitation programmes^26,48^ including the Clubhouse model (an accessible community space that acts as a social hub and promotes vocational rehabilitation),^56^ but not case management combined with individual rehabilitation^58^ or a digital psychosocial intervention based on principles of solution-focused therapy (DIALOG).^60^

**Table 3.** Studies measuring outcomes across multiple levels of the socio-ecological model with moderate-low risk of bias (excluding inpatient (psychiatric/unspecified) settings)

| Author | Year | Country | Design | Baseline total | Intervention | Platform | Individual-level outcome | Family-level outcome | Organisation-level outcome | Community-level outcome |
| --- | --- | --- | --- | --- | --- | --- | --- | --- | --- | --- |
| Guo et al<sup>26</sup> | 2010 | China | RCT (individual) | 1268 | Monthly manualised group psychosocial intervention for one year, including psycho-education, family intervention, skills training and cognitive-behavioural therapy | Specialist hospital out-patient | Adherence (SD), functioning (SD), mental health (NS/SD), QALYs (SD) |  | Service costs (NS/SD) | Employment (SD), Education (SD) |
| Ran et al<sup>27</sup> | 2015 | China | RCT (cluster) | 238 | Family psychoeducation once a month for nine months, family workshops every three months and crisis intervention when necessary | Community | Functioning (NS), mental health (NS/SD), mortality (NS) | Carer knowledge (SD), caregiving (SD) |  | Employment (SD) |
| Zhao et al<sup>32</sup> | 2013 | China | RCT (individual) | 105 | Monthly 40–60 minute online consultations or web-based psychoeducation for people living with psychosis and caregivers | Community | Mental health (SD), quality of life (SD), self-stigma (SD) |  |  | Social support for people living with psychosis (SD) |
| Tao et al<sup>35</sup> | 2021 | China | Meta-analysis | MA (1749 across included studies) | Mindfulness meditation (n=20 studies) | Other | Functioning (NS/SD), mental health (SD), personal recovery (SD), quality of life (NS) |  |  | Public stigma (SD), social support for people living with psychosis (NS) |
| Srinivasa Murthy et al<sup>43</sup> | 2005 | India | Uncontrolled before-after study | 100 | Community outreach comprising psychiatric assessment, outreach home visits, psychosocial care and family problem solving | Community | Functioning (SD), mental health (SD) | Carer burden (SD), household costs (SD), mental health of carer (SD),<br>physical health<br>of carer (SD) | Service costs (NS/ SD-) |  |
| Zhang et al<sup>44</sup> | 2013 | China | Controlled<br>clinical trial | 121 | Family intervention based<br>on psychoeducation and<br>mutual support | Community | Quality of<br>life (SD) |  |  | Social<br>support for<br>people living<br>with<br>psychosis<br>(SD) |
| Zhen et al<sup>45</sup> | 2012 | China | Controlled<br>clinical trial | 102 | Community service for<br>people living with<br>schizophrenia, including<br>regular training for staff | Community | Mental<br>health (SD) |  |  | Social<br>support for<br>people living<br>with<br>psychosis<br>(SD) |
| Sharif et al<sup>46</sup> | 2012 | Iran | Controlled<br>clinical trial | 70 | A group psycho-education<br>programme for caregivers<br>consisting of 10 90 minute<br>sessions held over five<br>weeks | Specialist hospital<br>out-patient | Mental<br>health (SD) | Carer burden<br>(SD) |  |  |
| Kulhara et al<sup>47</sup> | 2009 | India | RCT<br>(individual) | 152 | A structured<br>psychoeducation<br>intervention for caregivers<br>delivered during monthly<br>sessions across nine months | Hospital (not-specified) out-patient | Functioning<br>(SD), mental<br>health (SD) | Carer burden<br>(NS), carer<br>satisfaction<br>(SD) |  |  |
| Valencia et al<sup>48</sup> | 2010 | Mexico | Controlled<br>clinical trial | 83 | Psychosocial rehabilitation<br>comprising sessions with<br>PLWP, their caregivers and<br>combined sessions over one<br>year | Specialist hospital<br>out-patient | Adherence<br>(SD),<br>functioning<br>(SD), mental<br>health (SD) | Family<br>communication<br>(NS) | Engagement<br>of people<br>living with<br>psychosis<br>(SD) |  |
| Malakouti et al<sup>49</sup> | 2015 | Iran | RCT<br>(individual) | 180 | Monthly home visits by<br>either general practitioners<br>or nurses | Community | Functioning<br>(NS), mental<br>health (SD),<br>QALYs (SD) | Carer burden<br>(NS), carer<br>knowledge<br>(SD), carer<br>satisfaction<br>(NS) |  |  |
| Hasan et al<sup>50</sup> | 2015 | Jordan | RCT | 121 | Psychoeducation | Specialist hospital | Knowledge | Carer burden |  |  |
|  |  |  | (individual) |  | intervention for people living with psychosis and their caregivers, delivered every 2 weeks for 12 weeks | out-patient | (SD), mental health (SD) | (SD), carer knowledge (SD), carer quality of life (SD) |  |  |
| Koolae et al<sup>51</sup> | 2010 | Iran | RCT (individual) | 62 | Behavioural family management intervention for mothers of people living with psychosis | Hospital (not-specified) out-patient | Mental health (SD) | Carer burden (SD), family communication (SD) |  |  |
| Luo et al<sup>52</sup> | 2018 | China | RCT (individual) | 120 | Culturally adapted assertive community treatment involving two to three community/home visits per week and monthly family psychoeducation sessions for 12 months | Mixed hospital/community (e.g. home visits from a hospital) | Functioning (SD), mental health (SD) | Carer quality of life (SD), family functioning (SD) |  |  |
| Sungur et al<sup>53</sup> | 2011 | Turkey | RCT (individual) | 100 | Assertive case management comprising pharmacological and psychosocial treatment strategies | Mixed hospital/community (e.g. home visits from a hospital) | Functioning (SD), mental health (SD), quality of life (SD) | Carer mental health (SD) |  |  |
| Incedere et al<sup>54</sup> | 2019 | Turkey | Uncontrolled before-after study | 34 | Case management intervention | Mixed hospital/community (e.g. home visits from a hospital) | Adherence (SD), functioning (SD), mental health (SD) | Carer burden (SD) |  |  |
| Xu et al<sup>55</sup> | 2020 | China | Uncontrolled before-after study | 264 | 'Unlocking' restrained patients and transferring care to hospital or community services | Mixed hospital/community (e.g. home visits from a hospital) | Adherence (SD), functioning (SD), mental health (SD) | Carer burden (SD) |  |  |
| Yan et al<sup>56</sup> | 2021 | China | Meta-analysis | MA (682) | 'Clubhouse' model of psychiatric rehabilitation (n=7 studies) | Social care/community residential | Functioning (SD), mental health (SD), quality of life (SD) | Carer burden (SD) |  |  |
| Hanlon et al<sup>57</sup> | 2022 | Ethiopia | RCT (individual) | 329 | Task-shared care whereby primary health workers were trained to deliver mental health care integrated into their existing activities | Primary health care/ community health centre | Mental health (NS, time costs (NS) | Household costs (NS) | Service costs (NS/ SD) |  |
| Fang-Jian<sup>58</sup> | 2008 | China | Controlled clinical trial | 48 | Case management involving individual rehabilitation plans and regular follow-up for 11 months | Community | Functioning (SD), mental health (SD), mortality (SD) | Family functioning (NS) |  |  |
| Ofori-Atta<sup>41</sup> | 2018 | Ghana | RCT (individual) | 143 | Psychiatric care combined with treatment in a prayer camp | Community | Functioning (SD), mental health (SD/NS), quality of life (SD) |  | Human rights abuses (NS) |  |
| Hedge<sup>59</sup> | 2012 | India | RCT (individual) | 45 | Home-based cognitive retraining programme with instructions in a booklet | Community | Functioning (SD), mental health (SD) | Carer mental health (NS) |  |  |
| Asher<sup>31</sup> | 2022 | Ethiopia | RCT (cluster) | 332 | Community-based rehabilitation intervention lasting nine months and involving home visits by community health workers | Community | Adherence (SD), employment (SD-), functioning (SD/NS), mental health (SD/NS), mortality (NS) | Carer mental health (NS), human rights abuses by family (NS) |  | Public stigma (NS) |
| Chatterjee<sup>40</sup> | 2014 | India | RCT (individual) | 282 | Community-based rehabilitation intervention lasting 12 months and involving home visits by community health workers | Community | Adherence (SD), functioning (NS), knowledge (NS), mental health (NS) | Carer burden (NS) | Service costs (SD-) | Public stigma (NS) |
| Muhic<sup>29</sup> | 2022 | Bosnia and | RCT | 72 | Multi-family group | Specialist hospital | Mental |  |  | Social |
|  |  | Herzegovina | (individual) |  | meetings between people living with psychosis and one-two selected family members or friends, meeting once a month for six months | out-patient | health (SD/NS), quality of life (SD) |  |  | participation (NS) |
| Jovanovic<sup>60</sup> | 2022 | Bosnia and Herzegovina, Kosovo, Montenegro, North Macedonia, and Serbia | RCT (cluster) | 549 | DIALOG - a digital psychosocial intervention six times over 12 months (once per month for the first three months and then once every three months) | Hospital (not-specified) out-patient | Mental health (NS), quality of life (SD/NS) |  | Engagement of people living with psychosis (NS) |  |
| Meepring<sup>61</sup> | 2023 | Thailand | RCT (individual) | 106 | Physical health checks every three months up to five times in total | Specialist hospital out-patient | Physical health (SD), quality of life (NS) |  | Engagement of people living with psychosis (NS) |  |
| Fan<sup>38</sup> | 2022 | China | Uncontrolled before-after study | 167 | Community-based peer support | Community | Adherence (NS), functioning (SD), mental health (SD/NS), personal recovery (NS) | Carer mental health (NS), carer wellbeing (NS) |  |  |
| Jirapramukpitak<sup>28</sup> | 2022 | Thailand | Two group pre-post | 549 | Community health worker-led case management | Community | Mental health (SD-) |  |  | Participation in economic life (NS) |
| Sikira<sup>30</sup> | 2021 | Bosnia and Herzegovina | RCT (individual) | 65 | Volunteer befriending programme | Community | Mental health (SD), quality of life (SD) |  |  | Social participation (NS) |

\*Non-inferiority design
MA = meta-analysis; NS = non-statistically significant difference; RCT = randomised controlled trial; SD = statistically significant difference indicating potential benefit; SD- = statistically significant difference (unexpected direction)

Some studies of care models in the community e.g. psychiatric outreach, assertive case management and home visits by health professionals^43,49,52-55^ demonstrated significant differences in outcomes at multiple levels. However, three studies of care models delivered by community health workers did not show differences,^28,31,40^ nor did a study of collaborative care with faith healers,^41^ studies evaluating community peer support,^38^ or volunteer befriending.^30^ Otherwise, the tested interventions were heterogenous, with mindfulness meditation^35^ showing differences across multiple levels of the socio-ecological model, but not a physical health intervention^61^ or home-based cognitive retraining.^59^

Four studies reporting outcomes across multiple levels found significant differences in prioritised outcomes. Guo et al. (2010)^26^ studied a comprehensive group psychosocial intervention and linked individual outcomes (adherence, functioning, mental health) with community level outcomes (employment and access to education) at one year follow-up, but did not report possible mechanisms. Ran et al. (2015)^27^ tested family psychoeducation and found differences at the individual, family (knowledge and caregiving) and community level (employment) at 14-year follow-up. The authors speculated that improved caregiver knowledge and attitudes resulted in greater inclusion of the person with psychosis in family life (such as farming and household work), which may have improved their social functioning and ability to work. Zhao et al. (2013)^32^ reported that monthly online psychoeducation involving family resulted in enhanced perceived support, which predicted self-stigma in multiple regression analyses. Tao et al. (2021)^35^ found that no study in their meta-analysis investigated the mechanisms of mindfulness meditation quantitatively.

Only 12 of the 315 included studies reported economic evidence; of these, seven had moderate-low risk of bias. One was a partial evaluation study using cost analysis^43^ and six were full economic evaluation studies.^36,40,49,57,62,63^ All of the full economic evaluation studies adopted a healthcare sector perspective in costing but most included analyses from a societal perspective either as a complementary or sensitivity analysis. The outcome measures in the estimation of incremental cost effectiveness ratios (ICERs) varied between studies and evidence on cost-effectiveness was mixed. Two full economic evaluation studies reported that the respective interventions were cost-effective: the comprehensive group psychosocial intervention described by Guo et al. (2010)^26,62^ and a collaborative care approach.^36^ One evaluation found that DIALOG was not cost-effective.^63^ Two studies reported ICERs but did not compare these with a defined willingness-to-pay threshold or make a clear statement on cost-effectiveness.^40,49^ One study of integrated primary mental health care reported that the intervention was non-inferior and less costly than outpatient psychiatric nurse care augmented with community lay workers from a health-care perspective.^57^

### Risk of bias

Of the 315 studies included in this review, 22 were pilot or feasibility designs. Of the remaining 294 studies, 54% (159/294) were rated as being at high risk of bias, 28% (81/294) at moderate risk and 18% (53/294) at low risk of bias.

## Discussion

This systematic review is the most comprehensive synthesis to date of intervention research aimed at supporting the recovery of people living with psychosis and their family members in LMICs. We identified 310 individual intervention studies and a further five meta-analyses describing a diversity of approaches, far beyond the biomedical and risk or protection-focused care often provided. However, fewer than half of studies (excluding pilot and feasibility designs) were sufficiently rigorous to be included in our assessment of intervention effectiveness.

Our use of the socio-ecological model enabled mapping of a wide range of extracted outcomes across individual, family, organisational and community levels. Using this approach exposed a dominant focus of studies reporting individual-level mental health and to a lesser extent functioning outcomes, and a relative paucity of studies measuring the prioritised recovery-orientated outcomes of social inclusion, personal recovery, and reduced stigma and human rights abuses. Broadly, the evidence for effects on outcomes that matter most to people living with psychosis was limited; while over 90% of the 100 studies measuring individual mental health outcomes reported statistically significant effects, only around half of the 21 studies measuring prioritised outcomes reported statistically significant findings with generally small-to-modest effect sizes. This reflects the emphasis of research on clinical management of psychoses over social and personal recovery. It may also reflect an absence of valid measures of constructs prioritized by people living with psychosis that are sensitive to change. Comprehensive, contextualised and psychometrically valid measures for social inclusion and personal recovery were lacking in included studies. Instead, we relied largely on measurement of components of these constructs, such as employment, or scales that were comprehensive, if not contextualised to evaluate the evidence for effective interventions.

Plausibly lower effectiveness of interventions for priority recovery outcomes may also be attributable to the quality of development, adaptation and implementation of these complex interventions, and the extent to which people living with psychosis, and their families, were involved in their design. We found some evidence from China that multi-component interventions involving family, delivering psychoeducation and either additional skills and psychological therapies^26^ or crisis intervention^27^ resulted in significantly higher levels of employment than the comparator. There was also evidence of cost-effectiveness for these approaches.^26,62^ The small number of highly heterogeneous interventions examining impact on aspects of personal recovery makes it difficult to draw strong conclusions. The dearth of evidence for interventions able to reduce stigma and improve human rights protections is striking, although collaborative care model involving traditional faith healers at primary care level has shown some promise in reducing harmful treatment practices in Ghana and Nigeria.^36^

The socio-ecological model helps to conceptualise how intervention(s) at one level can impact different levels, and to pinpoint where and when multi-level and multi-component interventions are most needed. Again, we found evidence that family interventions can impact multiple levels, with Guo et al. (2010) positing a potential mechanism through which greater inclusion in family life is a stepping stone to wider inclusion.^26^ We found that models of psychosocial rehabilitation and psychiatric care in the community were effective over multiple levels but relied on specialist or trained health professionals.

Our systematic review exposes gaps in evidence for the mechanisms or necessary preconditions through which commonly measured individual-level outcomes, such as mental health and functioning, might link to improvements in more highly valued social inclusion outcomes. This could be a barrier to increasing the efficiency and impact of interventions. To address this challenge, trials require greater focus on programme theory and approaches such as causal mediation analysis alongside the traditional measurement of outcomes.^66^ In addition, theory-driven approaches such as realist evaluation, incorporating qualitative and quantitative data, have the potential to illuminate what works, how, for whom and in which contexts.^67^

We identified geographical inequity in the distribution of intervention studies for people living with psychosis, with most studies conducted in upper middle-income countries, particularly China, and mostly in urban settings. Little has changed in this regard since the first Lancet Series on Global Mental Health in 2007.^68^ Populations in low-income countries (mostly in Africa) and rural contexts continue to be underserved by research efforts, and even in these settings, most research is focused on intervention development and measuring symptom severity as the primary outcome. Heavy reliance on hospital platforms and mental health specialists for intervention delivery calls into question the extent to which available evidence can be applied or scaled in low-resource or rural settings.

Over half of included studies were at high risk of bias. Selection bias, confounding, lack of masking and other concerns meant that studies of potentially promising interventions could not be included in our effectiveness synthesis. There is a clear need to invest in strengthening capacity to conduct rigorous research to develop and evaluate complex interventions for psychosis in low-resource settings. Multi-country collaborations can play vital roles in driving research excellence and sustainable impacts in under-served populations. The United States National Institutes of Health mental health hubs^69^ and NIHR global health research units and groups^70^ have been influential and impactful, but USAID and UK Overseas Development Assistance cuts and resultant slashing of global health research funding pose major threats to mitigating these inequities.^71^

Our review has key strengths. We searched nine databases from 2001 to January 2024. We included studies published in all languages. We included all intervention study designs, capturing the breadth of research across diverse regions. We purposively included pilot, feasibility and small sample size or high-risk studies in our description of the intervention landscape, as these studies offer important information about the pipeline of intervention development across resource constrained, non-Western settings, but excluded these in our synthesis of evidence for effectiveness. We extracted a range of reported outcomes, balancing the need to prioritise evidence for recovery-orientated outcomes with synthesising the more commonly clinical outcomes from studies published over the last two decades. In addition, we provide a model through which intervention impacts can be understood at different levels: individual, family, organisation and community. We considered that the socio-ecological approach would be important in LMICs where contexts vary considerably, services have limited reach, and where family and community life may assume greater importance to recovery.

However, this approach may also contribute a helpful perspective to evaluating impacts of interventions in high-income countries.

This systematic review also has some limitations. Our focus on enrolling people living with psychosis and/or family members as participants, meant that we did not synthesise evidence for interventions targeting other groups (e.g. health workers or community members). The process of mapping domains of recovery to levels of the socio-ecological model was subjective. Domains such as treatment adherence could be re-conceptualised as a service level outcome, rather than an individual level outcome. Furthermore, our conceptualisation did not take into account wider politico-economic contexts, despite their importance for determining social inclusion across population groups.^72^ For non-prioritised outcomes, we categorised effectiveness evidence as statistically significant or non-significant at the first time point. As a result, our synthesis cannot comment on the intervention effect sizes across all outcome types (in order to make judgements on what is meaningful to an individual) or the sustainability of impacts over time.

In conclusion, we systematically mapped the evidence landscape from two decades of research on interventions for people living with psychosis and their family in LMICs and revealed critical gaps and imbalances in the global evidence base. Gaps in existing evidence include neglect of recovery-orientated outcomes valued by people living with psychosis, the predominance of individual-level outcomes, failure to consider the person living with psychosis within their wider context, and missed opportunities to investigate intervention mechanisms and cost-effectiveness. There is some evidence in support of specialist-delivered comprehensive interventions involving family, psychosocial rehabilitation and care close to home, but many intervention types and delivery agents have not been adequately tested, especially in LICs and rural settings. Involvement of people living with psychosis and their families in the co-production of optimised interventions and the use of contextualised measures of recovery are required to direct the focus of future psychosis intervention research in low-income and middle-income settings.^11^

## Data Availability

All data produced in the present study are available upon reasonable request to the authors

## Contributions

SB, CH, AKM, SC, BC, NJ, SC, JE conceived the study.

AB, CH, AKM, SC, BC, NJ, SC, JE, AM, RG, SX and EW designed the study.

SB, RG, WF and AW conducted the literature search and SB, LM, SX, RG, CJ, WF, AM, HW, RCK, EW and CH independently screened papers and extracted data. EA extracted cost data.

SB, LM, SX, RG, WF, AM, HW, RCK, CH and AKM conceived the process for the narrative synthesis.

SB conducted the narrative synthesis and data interpretation with input from CH and AKM. EA conducted the assessment for evidence of cost-effectiveness.

SB wrote the first draft of the report with contributions from CH and AKM. All authors contributed to subsequent drafts and approved the final report. All authors had full access to all the data in the study and had final responsibility for the decision to submit for publication.

## Declarations

We declare no competing interests.

## Data sharing

All data underlying the findings presented in the main manuscript are included in the supplementary files.

## Acknowledgment

Dr Sahar Fazeli and Dr Passachol Makprakhon are gratefully acknowledged for their contribution to translation of papers published in Persian and Thai.

SB was employed as an NIHR-funded Clinical Academic Fellow in General Adult Psychiatry and later as a Wellcome Trust funded CREATE PhD Fellow during this study.

RCK was employed as an NIHR-funded Clinical Lecturer in General Adult Psychiatry during this study. Her research is supported by an Academy of Medical Sciences Starter Grant for Clinical Lecturers (SGL030\1047).

CH receives funding support from the National Institute for Health and Care Research (NIHR) through the NIHR Global Health Research Group on Homelessness and Mental Health in Africa (NIHR134325) using UK aid from the UK Government. The views expressed in this publication are those of the authors and not necessarily those of the NIHR or the Department of Health and Social Care. CH and WF receive support from Wellcome grants 222154/Z20/Z. CH is also supported through Wellcome grant 223615/Z/21/Z.

For the purposes of open access, the author has applied a Creative Commons Attribution (CC BY) licence to any Accepted Author Manuscript version arising from this submission

