## Supplementary Files for "Effective interventions to support recovery of people with psychosis and their families across socio-ecological levels in low-income and middle-income countries: a systematic review"

**Supplementary Materials**

### **Supplementary file 1: Minor deviations from the protocol**

1. Review title

• Old version: Effective and cost-effective interventions to support recovery of people with lived experience of psychosis and their caregivers in low- and middle-income countries (LMICs): a systematic review

• New version: Effective interventions to support recovery of people with psychosis and their families across socio-ecological levels in low-income and middle-income countries: a systematic review

2. Searches

Addition of Tufts Medical Canter Cost-Effectiveness Analysis (CEA) Registry

3. Study design

• Naturalistic studies were not considered a separate category of design

• Meta-analyses of a focused intervention (e.g. Mindfulness meditation) in a homogenous population (e.g. single country) were included as a whole, without extraction of data from individual studies

4. Population

• Clarification: Studies of bipolar disorder required a psychosis specifier.

• Clarification: Studies of long-term institutional care as an intervention would not be included, however studies of interventions taking place within inpatient settings would be included.

5. Main outcomes

The primary outcome of interest was defined as social inclusion of people with psychosis. In the drafting of the manuscript we revised our primary outcomes of interest to also include outcomes relating to personal recovery, stigma, discrimination and human rights abuses.

6. Risk of bias assessment

We used the Effective Public Health Practice Project (EPHPP) tool instead of the Cochrane risk of bias assessment tool. The CHEERS Checklist was not used to assess risk of bias in included cost-effectiveness studies, as they had been rated by the EPHPP. The AMSTAR-2 tool was used to evaluate the risk of bias of included meta-analyses.

### **Supplementary file 2: Search terms (PubMed)**

The following Pubmed search terms have been re-organised for readability using generative AI (ChatGPT version 5.0) and has been cross-checked against the Pubmed search log for accuracy. The full original search terms for all databases can be provided on request.

Search terms were combined as follows: CONDITION **AND** INTERVENTION **AND** SETTING **AND** STUDY DESIGN

1. CONDITION

[Psychosis terms]

("psychotic"[Title/Abstract] OR

"psychosis"[Title/Abstract] OR

"psychoses"[Title/Abstract])

[Disorders]

OR "psychotic disorders"[MeSH Terms] OR

"affective disorders, psychotic"[MeSH Terms] OR

"affective disorders, psychotic"[MeSH Terms] OR

"schizophrenia"[MeSH Terms] OR

"bipolar and related disorders"[MeSH Terms] OR

"perceptual disorder*"[Title/Abstract]

[Perinatal/childbirth psychosis]

OR (("perinatal"[All Fields] OR

"perinatally"[All Fields] OR

AND "psychosis"[Title/Abstract])

OR "puerperal psychosis"[Title/Abstract]

OR (("psychotic disorders"[MeSH Terms] OR

("psychotic"[All Fields] AND "disorders"[All Fields]) OR

"psychotic disorders"[All Fields] OR

"psychosis"[All Fields])

AND "after childbirth"[Title/Abstract])

[Symptoms]

OR "hallucinations"[MeSH Terms] OR

"delusions"[MeSH Terms] OR

"hear* voice*"[Title/Abstract] OR

"hearing voices"[Title/Abstract] OR

"voice hearing"[Title/Abstract] OR

"hallucinat*"[Title/Abstract] OR

"delusion*"[Title/Abstract] OR

"paranoi*"[Title/Abstract]

[Severe mental illness / psychosocial disability]

OR "severe mental"[Title/Abstract] OR

"serious mental"[Title/Abstract] OR

"psychosocial disability"[Title/Abstract] OR

"psychosocial disabilities"[Title/Abstract] OR

"psychosocial disorders"[Title/Abstract]

2. INTERVENTION

[Psychological therapies and education]

("psychotherapy"[MeSH Terms] OR

"counseling"[MeSH Terms] OR

"telemedicine"[MeSH Terms] OR

("telehealth"[Title/Abstract] OR "mhealth"[Title/Abstract] OR "telepsy*"[Title/Abstract]) OR

"health education"[MeSH Terms] OR

"patient education as topic"[MeSH Terms] OR

("psychoeducation*"[Title/Abstract] OR "psycho education*"[Title/Abstract]))

[Family / caregiver education and support]

OR (("patient*"[Title/Abstract] OR "caregiver*"[Title/Abstract] OR "care giver*"[Title/Abstract] OR "carer*"[Title/Abstract] OR "family"[Title/Abstract] OR "families"[Title/Abstract])

AND ("education"[Title/Abstract] OR "advice"[Title/Abstract] OR "information"[Title/Abstract] OR "Training"[Title/Abstract] OR "support"[Title/Abstract] OR "intervention"[Title/Abstract] OR "livelihood"[Title/Abstract]))

OR (("family"[Title/Abstract] OR "families"[Title/Abstract] OR "caregiver"[Title/Abstract] OR "caregivers"[Title/Abstract] OR "care-giver"[Title/Abstract] OR "care"[Title/Abstract]) AND "burden"[Title/Abstract])

[Medication adherence and management]

OR ("patient compliance"[MeSH Terms] OR "medication adherence"[MeSH Terms])

OR ("patient"[Title/Abstract] AND ("compliance"[Title/Abstract] OR "concordance"[Title/Abstract] OR "adherence"[Title/Abstract]))

OR (("antipsychotic*"[Title/Abstract] OR "anti psychotic*"[Title/Abstract] OR "medication"[Title/Abstract])

AND ("compliance"[Title/Abstract] OR "concordance"[Title/Abstract] OR "adherence"[Title/Abstract] OR "non-compliance"[Title/Abstract] OR "non-concordance"[Title/Abstract] OR "non-adherence"[Title/Abstract]))

OR (("adherence"[Title/Abstract] OR "compliance"[Title/Abstract] OR "medication"[Title/Abstract])

AND ("support"[Title/Abstract] OR "therapy"[Title/Abstract] OR "education"[Title/Abstract] OR "Training"[Title/Abstract] OR "advice"[Title/Abstract] OR "information"[Title/Abstract] OR "intervention"[Title/Abstract]))

OR (("antipsychotic*"[Title/Abstract] OR "anti psychotic*"[Title/Abstract] OR "medication"[Title/Abstract])

AND ("choice"[Title/Abstract] OR "decision"[Title/Abstract]))

OR ("medication therapy management"[MeSH Terms] OR "pharmacists"[MeSH Terms] OR "polypharmacy"[MeSH Terms])

OR ("side-effect"[Title/Abstract] OR "adverse reaction"[Title/Abstract] OR "contraindicat*"[Title/Abstract])

OR "medication discontinuation"[Title/Abstract]

OR ("delayed action preparation"[Title/Abstract] OR "depot"[Title/Abstract] OR "long-acting"[Title/Abstract])

[Decision-making and directives]

OR ("decision-making"[Title/Abstract] OR "decision support techniques"[Title/Abstract] OR "advance directives"[Title/Abstract])

OR ("decision-making"[MeSH Terms] OR "advance directives"[MeSH Terms])

[Rehabilitation, social inclusion and community-based care]

OR ("social inclusion"[MeSH Terms] OR "social inclusion"[Title/Abstract])

OR ("Rehabilitation"[MeSH Terms] OR "rehabilitation centers"[MeSH Terms] OR "rehabilitation nursing"[MeSH Terms] OR "psychiatric rehabilitation"[MeSH Terms] OR "hospitals, rehabilitation"[MeSH Terms] OR "rehabilitation, vocational"[MeSH Terms] OR "employment, supported"[MeSH Terms] OR "social adjustment"[MeSH Terms] OR "cooperative behavior"[MeSH Terms] OR "interpersonal relations"[MeSH Terms])

OR ("community based rehabilitation"[Title/Abstract] OR "CBR"[Title/Abstract])

OR ("communit*"[Title/Abstract] AND ("vocational training"[Title/Abstract] OR "apprenticeship*"[Title/Abstract] OR (("employability"[All Fields] OR "employment"[MeSH Terms] OR "employment"[All Fields]) AND "placement service*"[Title/Abstract]) OR "support network*"[Title/Abstract] OR "self employ*"[Title/Abstract] OR "supported employ*"[Title/Abstract] OR "social service*"[Title/Abstract] OR "social work*"[Title/Abstract]))

OR ("communit*"[Title/Abstract] AND ("personal assistance"[Title/Abstract] OR "personal assistant*"[Title/Abstract] OR "individual support*"[Title/Abstract] OR (("disabilities"[All Fields] OR "persons with disabilities"[MeSH Terms] OR "disabled"[All Fields]) AND ("organisation"[Title/Abstract] OR "organization"[Title/Abstract]))))

OR ("communit*"[Title/Abstract] AND ("empower*"[Title/Abstract] OR "awareness campaign*"[Title/Abstract] OR "self-advocacy"[Title/Abstract] OR "self help group*"[Title/Abstract] OR "support group*"[Title/Abstract] OR "women* group*"[Title/Abstract] OR "development group*"[Title/Abstract]))

OR "self-management"[Title/Abstract]

OR ("community"[Title/Abstract] AND "inclusi*"[Title/Abstract] AND ("health"[Title/Abstract] OR "education"[Title/Abstract] OR "hous*"[Title/Abstract] OR "Social"[Title/Abstract] OR "justice"[Title/Abstract] OR "empower*"[Title/Abstract]))

OR ("Rehabilitation"[Title/Abstract] AND ("vocational"[Title/Abstract] OR "Social"[Title/Abstract] OR "personal"[Title/Abstract]))

OR ("Training"[Title/Abstract] AND ("life skill*"[Title/Abstract] OR "social skill*"[Title/Abstract] OR "personal skill*"[Title/Abstract] OR "interpersonal skill*"[Title/Abstract] OR "interpersonal"[Title/Abstract]))

OR (("Psychosocial"[Title/Abstract] OR "psycho-social"[Title/Abstract] OR "Social"[Title/Abstract] OR "psychological"[Title/Abstract] OR "psychiatric"[Title/Abstract] OR "PSR"[Title/Abstract] OR "vocational"[Title/Abstract] OR "occupational"[Title/Abstract])

AND ("intervention*"[Title/Abstract] OR "support"[Title/Abstract] OR "Rehabilitation"[Title/Abstract]))

OR ("Recovery"[Title/Abstract] OR "recovery model"[Title/Abstract] OR "recovery approach"[Title/Abstract] OR "social recovery"[Title/Abstract])

[Sustainable livelihoods and economic supports]

OR (("sustainable livelihood*"[Title/Abstract] OR "livelihood*"[Title/Abstract]) AND ("intervention"[Title/Abstract] OR "support"[Title/Abstract]))

OR ("education"[Title/Abstract] AND ("support"[Title/Abstract] OR "intervention*"[Title/Abstract] OR "inclusion"[Title/Abstract]))

OR ("microfinance"[Title/Abstract] OR "financial support"[MeSH Terms])

OR "cash transfer"[Title/Abstract]

OR ("social security"[MeSH Terms] OR "disability benefit"[Title/Abstract] OR "poverty reduction"[Title/Abstract] OR "food assistance"[MeSH Terms])

[Health promotion and physical comorbidities]

OR "health promotion"[MeSH Terms]

OR ("health"[Title/Abstract] AND ("promotion"[Title/Abstract] OR "advice"[Title/Abstract] OR "information"[Title/Abstract] OR "Training"[Title/Abstract] OR "support"[Title/Abstract]))

OR ("life style"[MeSH Terms] OR "self care"[Title/Abstract])

OR ("wellbeing"[Title/Abstract] OR "well-being"[Title/Abstract])

OR ("family planning"[Title/Abstract] OR "family planning services"[MeSH Terms] OR "mass screening"[MeSH Terms] OR "nutrition therapy"[MeSH Terms] OR "nutrition assessment"[MeSH Terms] OR "malnutrition"[MeSH Terms])

OR ("co-morbidity"[Title/Abstract] OR "comorbidity"[Title/Abstract])

OR ("metabolic syndrome"[MeSH Terms] OR "syphilis"[MeSH Terms] OR "hypertension"[MeSH Terms] OR "diabetes mellitus"[MeSH Terms] OR "hypoglycaemic agents"[Title/Abstract] OR "insulin"[MeSH Terms] OR "body weight"[MeSH Terms] OR "heart diseases"[MeSH Terms] OR "cardiovascular diseases"[MeSH Terms] OR "neoplasms"[MeSH Terms] OR "pulmonary disease, chronic obstructive"[MeSH Terms] OR "neglected diseases"[MeSH Terms])

OR "neglected tropical dis*"[Title/Abstract]

[Peer and social support]

OR ("social support"[MeSH Terms] OR ("self help groups"[Title/Abstract]))

OR (("Social"[Title/Abstract] OR "peer*"[Title/Abstract] OR "peer-led"[Title/Abstract] OR "self-help"[Title/Abstract] OR "community"[Title/Abstract] OR "cooperative"[Title/Abstract] OR "co-operative"[Title/Abstract])

AND ("group"[Title/Abstract] OR "support"[Title/Abstract] OR "support group"[Title/Abstract]))

[Participation, stigma and rights]

OR ("patient involvement"[Title/Abstract] OR "patient participation"[MeSH Terms])

OR ("community participation"[MeSH Terms] OR "service user involvement"[Title/Abstract] OR "social stigma"[MeSH Terms] OR "stigma reduction"[Title/Abstract] OR "human rights"[MeSH Terms])

OR ("exploit"[Title/Abstract] OR "discriminat*"[Title/Abstract] OR "abuse"[Title/Abstract] OR "rights"[Title/Abstract] OR "empower"[Title/Abstract] OR "self-advocacy"[Title/Abstract])

OR ("care"[Title/Abstract] AND ("collaborate"[Title/Abstract] OR "community"[Title/Abstract] OR "community-based"[Title/Abstract]))

OR ("outreach"[Title/Abstract] AND ("service*"[Title/Abstract] OR "care"[Title/Abstract] OR "intervention*"[Title/Abstract] OR "program*"[Title/Abstract]))

[Community and health service delivery]

OR ("community health services"[MeSH Terms] OR "home care services"[MeSH Terms] OR "home nursing"[MeSH Terms] OR "community health nursing"[MeSH Terms] OR "community networks"[MeSH Terms] OR "community mental health services"[MeSH Terms] OR "patient care team"[MeSH Terms] OR "nursing, team"[MeSH Terms] OR "social work"[MeSH Terms] OR "community health centers"[MeSH Terms] OR "community health workers"[MeSH Terms] OR "outpatients"[MeSH Terms] OR "ambulatory care facilities"[MeSH Terms] OR "outpatient clinics, hospital"[MeSH Terms] OR "ambulatory care"[MeSH Terms] OR "allied health personnel"[MeSH Terms] OR "psychiatric aides"[MeSH Terms])

OR ("lay worker"[Title/Abstract] OR "early intervention"[Title/Abstract] OR "first contact"[Title/Abstract] OR "first episode"[Title/Abstract] OR "case find*"[Title/Abstract] OR "key informant"[Title/Abstract] OR "assertive outreach team"[Title/Abstract] OR ("accident"[Title/Abstract] AND "emergency"[Title/Abstract]) OR "seclusion"[Title/Abstract] OR "coercion"[Title/Abstract] OR "assertive community team"[Title/Abstract] OR "assertive community treatment"[Title/Abstract] OR "emergency department"[Title/Abstract] OR "community mental health"[Title/Abstract])

OR ("case management"[MeSH Terms] OR "crisis intervention"[MeSH Terms])

OR ("hospitalization"[MeSH Terms] OR "coercion"[MeSH Terms] OR "hospitals, psychiatric"[MeSH Terms] OR "emergency service, hospital"[MeSH Terms] OR "emergency services, psychiatric"[MeSH Terms] OR "patient admission"[MeSH Terms] OR "patient discharge"[MeSH Terms] OR "mental health services"[MeSH Terms])

[Traditional, complementary and faith-based care]

OR ("medicine, african traditional"[MeSH Terms] OR "faith healing"[MeSH Terms] OR "complementary therapies"[MeSH Terms] OR "spiritual therapies"[MeSH Terms] OR ("alternative therapy"[Title/Abstract] OR "complementary medicine"[Title/Abstract]))

[Forensic, legal and coercive settings]

OR ("prisons"[MeSH Terms] OR "police"[MeSH Terms] OR "forensic psychiatry"[MeSH Terms] OR "criminals"[MeSH Terms] OR "prisoners"[MeSH Terms] OR "violence"[MeSH Terms] OR "aggression"[MeSH Terms] OR "antisocial personality disorder"[MeSH Terms] OR "crime victims"[MeSH Terms])

OR ("offender"[Title/Abstract] OR "criminal"[Title/Abstract] OR "prisoner"[Title/Abstract] OR "inmate"[Title/Abstract] OR "violent"[Title/Abstract] OR "victim"[Title/Abstract] OR "exploit"[Title/Abstract])

OR ("commitment of persons with psychiatric disorders"[MeSH Terms] OR "involuntary treatment"[MeSH Terms])

OR ("protection"[Title/Abstract] OR "revolving door"[Title/Abstract] OR ("community treatment order"[Title/Abstract] OR "cto"[Title/Abstract] OR "forced treatment"[Title/Abstract] OR "mental health act"[Title/Abstract] OR "mental health legislation"[Title/Abstract] OR "mental health law"[Title/Abstract]))

[Other populations and settings]

OR ("dual diagnosis"[Title/Abstract] OR "diagnosis, dual psychiatry"[MeSH Terms])

OR "mental competency"[MeSH Terms]

OR ("readmission"[Title/Abstract] OR "complex needs"[Title/Abstract])

OR ("alcoholism"[MeSH Terms] OR "alcohol"[Title/Abstract])

OR ("drug abuse"[Title/Abstract] OR "drug misuse"[Title/Abstract] OR "drug addiction"[Title/Abstract] OR "drug dependence"[Title/Abstract] OR "substance use disorder"[Title/Abstract] OR "substance use"[Title/Abstract] OR "substance dependency"[Title/Abstract] OR "substance dependence"[Title/Abstract])

OR ("substance related disorders"[MeSH Terms] OR "substance abuse treatment centers"[MeSH Terms])

OR ("pregnancy"[MeSH Terms] OR "pregnancy complications"[MeSH Terms] OR "prenatal care"[MeSH Terms] OR "postpartum period"[MeSH Terms])

OR ("pregnant"[Title/Abstract] OR "pregnancy"[Title/Abstract] OR "puerperium"[Title/Abstract] OR "puerperal"[Title/Abstract] OR "perinatal"[Title/Abstract] OR "prenatal"[Title/Abstract] OR "antenatal"[Title/Abstract] OR "postpartum"[Title/Abstract] OR "postnatal"[Title/Abstract])

OR ("neurocognitive disorders"[MeSH Terms] OR "neurodevelopmental disorders"[MeSH Terms] OR "autistic disorder"[MeSH Terms] OR "autism spectrum disorder"[MeSH Terms] OR "learning disabilities"[MeSH Terms] OR "attention deficit disorder with hyperactivity"[MeSH Terms] OR "hearing loss"[MeSH Terms] OR "vision disorders"[MeSH Terms] OR "ataxia"[MeSH Terms] OR "psychomotor disorders"[MeSH Terms] OR "cerebral palsy"[MeSH Terms])

OR ("mental retardation"[Title/Abstract] OR "cognitive impair*"[Title/Abstract] OR "mental impair*"[Title/Abstract] OR "mental disability"[Title/Abstract] OR "intellectual impair*"[Title/Abstract] OR "intellectual disorder"[Title/Abstract] OR "learning difficulty"[Title/Abstract])

OR ("behaviour problem"[Title/Abstract] OR "behavior problem"[Title/Abstract] OR "behaviour* disorder"[Title/Abstract] OR "behavior* disorder"[Title/Abstract] OR ("attention deficit"[Title/Abstract] AND "hyperactivity disorder"[Title/Abstract]) OR ("hyperkinetic"[Title/Abstract] OR "adhd"[Title/Abstract] OR "autism"[Title/Abstract] OR "language difficulty"[Title/Abstract] OR "hearing impairment"[Title/Abstract] OR "visual impairment"[Title/Abstract] OR "ataxia"[Title/Abstract] OR "motor impairment"[Title/Abstract] OR "psychomotor disorder"[Title/Abstract] OR "cerebral palsy"[Title/Abstract]))

OR ("treatment-resistant"[Title/Abstract] OR "treatment-refractory"[Title/Abstract])

OR ("homeless"[Title/Abstract] OR "homeless youth"[MeSH Terms])

OR ("vagabond"[Title/Abstract] OR "destitut*"[Title/Abstract] OR "street people"[Title/Abstract] OR "street person"[Title/Abstract] OR "abandonment"[Title/Abstract] OR "neglect"[Title/Abstract] OR "exploit*"[Title/Abstract])

OR "restraint, physical"[MeSH Terms]

[Health system, quality and policy]

OR ("quality rights"[Title/Abstract] OR "health system"[Title/Abstract] OR "detection"[Title/Abstract] OR "identification"[Title/Abstract] OR "engagement"[Title/Abstract] OR "chronic care"[Title/Abstract] OR "continuing care"[Title/Abstract])

OR ("patient safety"[MeSH Terms] OR "quality improvement"[MeSH Terms] OR "health services accessibility"[MeSH Terms] OR "health plan implementation"[MeSH Terms] OR "patient care planning"[MeSH Terms] OR "delivery of health care"[MeSH Terms]

3. SETTING

[MeSH heading]

("developing countries"[MeSH Terms])

[General descriptors: country/population]

OR (("developing"[Title/Abstract] OR "less developed"[Title/Abstract] OR "under developed"[Title/Abstract] OR "underdeveloped"[Title/Abstract] OR "middle income"[Title/Abstract] OR "low income"[Title/Abstract] OR "lower income"[Title/Abstract])

AND ("countr*"[Title/Abstract] OR "nation*"[Title/Abstract] OR "population*"[Title/Abstract] OR "world"[Title/Abstract]))

[General descriptors: economy/economies]

OR (("transition*"[Title/Abstract] OR "developing"[Title/Abstract] OR "less developed"[Title/Abstract] OR "lesser developed"[Title/Abstract] OR "under developed"[Title/Abstract] OR "underdeveloped"[Title/Abstract] OR "middle income"[Title/Abstract] OR "low income"[Title/Abstract] OR "lower income"[Title/Abstract])

AND ("economy"[Title/Abstract] OR "economies"[Title/Abstract]))

[Economic indicators and acronyms]

OR (("low"[Title/Abstract] AND ("gdp"[Title/Abstract] OR "gnp"[Title/Abstract] OR "gross domestic"[Title/Abstract] OR "gross national"[Title/Abstract]))

OR ("lmic"[Title/Abstract] OR "lmics"[Title/Abstract] OR "lamics"[Title/Abstract] OR "lamic"[Title/Abstract] OR "third world"[Title/Abstract] OR "lami countries"[Title/Abstract] OR "lami country"[Title/Abstract])

OR ("transitional country"[Title/Abstract] OR "transitional countries"[Title/Abstract]))

[Alternative descriptor]

OR "global south"[Title/Abstract]

[Country names]

OR ("Afghanistan"[Title/Abstract]

OR "Albania"[Title/Abstract]

OR "Algeria"[Title/Abstract]

OR "American Samoa"[Title/Abstract]

OR "Angola"[Title/Abstract]

OR "Argentina"[Title/Abstract]

OR "Armenia"[Title/Abstract]

OR "Azerbaijan"[Title/Abstract]

OR "Bangladesh"[Title/Abstract]

OR "Belarus"[Title/Abstract]

OR "Belize"[Title/Abstract]

OR "Benin"[Title/Abstract]

OR "Bhutan"[Title/Abstract]

OR "Bolivia"[Title/Abstract]

OR ("Bosnia"[Title/Abstract] AND "Herzegovina"[Title/Abstract])

OR "Botswana"[Title/Abstract]

OR "Brazil"[Title/Abstract]

OR "Bulgaria"[Title/Abstract]

OR "Burkina Faso"[Title/Abstract]

OR "Burundi"[Title/Abstract]

OR "Cabo Verde"[Title/Abstract]

OR "Cambodia"[Title/Abstract]

OR "Cameroon"[Title/Abstract]

OR "Central African Republic"[Title/Abstract]

OR "Chad"[Title/Abstract]

OR "Chile"[Title/Abstract]

OR "China"[Title/Abstract]

OR "Colombia"[Title/Abstract]

OR "Comoros"[Title/Abstract]

OR "Congo"[Title/Abstract]

OR "Costa Rica"[Title/Abstract]

OR "Cote d Ivoire"[Title/Abstract]

OR "Cuba"[Title/Abstract]

OR "Democratic People’s Republic of Korea"[Title/Abstract]

OR "Democratic Republic of the Congo"[Title/Abstract]

OR "Djibouti"[Title/Abstract]

OR "Dominica"[Title/Abstract]

OR "Dominican Republic"[Title/Abstract]

OR "Ecuador"[Title/Abstract]

OR "Egypt"[Title/Abstract]

OR "El Salvador"[Title/Abstract]

OR "Equatorial Guinea"[Title/Abstract]

OR "Eritrea"[Title/Abstract]

OR "Eswatini"[Title/Abstract]

OR "Ethiopia"[Title/Abstract]

OR "Fiji"[Title/Abstract]

OR "Gabon"[Title/Abstract]

OR "Gambia"[Title/Abstract]

OR "Ghana"[Title/Abstract]

OR "Georgia"[Title/Abstract]

OR "Grenada"[Title/Abstract]

OR "Guatemala"[Title/Abstract]

OR "Guinea"[Title/Abstract]

OR "Guinea-Bissau"[Title/Abstract]

OR "Guyana"[Title/Abstract]

OR "Haiti"[Title/Abstract]

OR "Honduras"[Title/Abstract]

OR "India"[Title/Abstract]

OR "Indonesia"[Title/Abstract]

OR "Iran"[Title/Abstract]

OR "Islamic Republic of Iran"[Title/Abstract]

OR "Iraq"[Title/Abstract]

OR "Jamaica"[Title/Abstract]

OR "Jordan"[Title/Abstract]

OR "Kazakhstan"[Title/Abstract]

OR "Kenya"[Title/Abstract]

OR "Kiribati"[Title/Abstract]

OR "Kosovo"[Title/Abstract]

OR "Kyrgyz Republic"[Title/Abstract]

OR "Lao People’s Democratic Republic"[Title/Abstract]

OR "Lebanon"[Title/Abstract]

OR "Lesotho"[Title/Abstract]

OR "Liberia"[Title/Abstract]

OR "Libya"[Title/Abstract]

OR "Madagascar"[Title/Abstract]

OR "Malawi"[Title/Abstract]

OR "Malaysia"[Title/Abstract]

OR "Maldives"[Title/Abstract]

OR "Mali"[Title/Abstract]

OR "Marshall Islands"[Title/Abstract]

OR "Mauritania"[Title/Abstract]

OR "Mauritius"[Title/Abstract]

OR "Mexico"[Title/Abstract]

OR "Micronesia"[Title/Abstract]

OR "Federated States of Micronesia"[Title/Abstract]

OR "Moldova"[Title/Abstract]

OR "Mongolia"[Title/Abstract]

OR "Montenegro"[Title/Abstract]

OR "Morocco"[Title/Abstract]

OR "Mozambique"[Title/Abstract]

OR "Myanmar"[Title/Abstract]

OR "Namibia"[Title/Abstract]

OR "Nauru"[Title/Abstract]

OR "Nepal"[Title/Abstract]

OR "Nicaragua"[Title/Abstract]

OR "Niger"[Title/Abstract]

OR "Nigeria"[Title/Abstract]

OR "North Macedonia"[Title/Abstract]

OR "Pakistan"[Title/Abstract]

OR "Palau"[Title/Abstract]

OR "Panama"[Title/Abstract]

OR "Papua New Guinea"[Title/Abstract]

OR "Paraguay"[Title/Abstract]

OR "Peru"[Title/Abstract]

OR "Philippines"[Title/Abstract]

OR "Rumania"[Title/Abstract]

OR "Romania"[Title/Abstract]

OR "Russian Federation"[Title/Abstract]

OR "Russia"[Title/Abstract]

OR "Rwanda"[Title/Abstract]

OR "Saint Lucia"[Title/Abstract]

OR "Samoa"[Title/Abstract]

OR "Senegal"[Title/Abstract]

OR "Serbia"[Title/Abstract]

OR "Seychelles"[Title/Abstract]

OR "Sierra Leone"[Title/Abstract]

OR "Solomon Islands"[Title/Abstract]

OR "Somalia"[Title/Abstract]

OR "South Africa"[Title/Abstract]

OR "South Sudan"[Title/Abstract]

OR "Sri Lanka"[Title/Abstract]

OR "State of Palestine"[Title/Abstract]

OR "Sudan"[Title/Abstract]

OR "Suriname"[Title/Abstract]

OR "Syrian Arab Republic"[Title/Abstract]

OR "Tajikistan"[Title/Abstract]

OR "Tanzania"[Title/Abstract]

OR "Thailand"[Title/Abstract]

OR "Timor Leste"[Title/Abstract]

OR "Togo"[Title/Abstract]

OR "Tonga"[Title/Abstract]

OR "Tunisia"[Title/Abstract]

OR "Turkey"[Title/Abstract]

OR "Turkmenistan"[Title/Abstract]

OR "Tuvalu"[Title/Abstract]

OR "Uganda"[Title/Abstract]

OR "Ukraine"[Title/Abstract]

OR "United Republic of Tanzania"[Title/Abstract]

OR "Uruguay"[Title/Abstract]

OR "Uzbekistan"[Title/Abstract]

OR "Vanuatu"[Title/Abstract]

OR "Venezuela"[Title/Abstract]

OR "Viet Nam"[Title/Abstract]

OR "Vietnam"[Title/Abstract]

OR ("West Bank"[Title/Abstract] AND "Gaza"[Title/Abstract])

OR "Yemen"[Title/Abstract]

OR "Zambia"[Title/Abstract]

OR "Zimbabwe"[Title/Abstract]

OR ("Antigua"[Title/Abstract] AND "Barbuda"[Title/Abstract])

OR ("Saint Kitts"[Title/Abstract] AND "Nevis"[Title/Abstract])

OR ("Saint Vincent"[Title/Abstract] AND "the Grenadines"[Title/Abstract])

OR ("Sao Tome"[Title/Abstract] AND "Principe"[Title/Abstract]))

4. STUDY DESIGN

[Before/after and quasi-experimental designs]

("before-after"[Title/Abstract] OR

"interrupted time series analysis"[MeSH Terms] OR

"pre post evaluation"[Title/Abstract] OR

"cohort studies"[MeSH Terms] OR

"naturalistic observation"[Title/Abstract] OR

"quasi-experimental"[Title/Abstract] OR

"non random*"[Title/Abstract])

[Randomised and controlled trials]

OR "randomized controlled trials as topic"[MeSH Terms] OR

"controlled clinical trials as topic"[MeSH Terms] OR

"random*"[Title/Abstract] OR

"randomization"[Title/Abstract] OR

"intermethod comparison"[Title/Abstract] OR

"placebo"[Title/Abstract] OR

("compare"[Title/Abstract] OR "compared"[Title/Abstract] OR "comparison"[Title/Abstract]) OR

(("Evaluated"[Title/Abstract] OR "evaluate"[Title/Abstract] OR "evaluating"[Title/Abstract] OR "assessed"[Title/Abstract] OR "assess"[Title/Abstract])

AND ("compare"[Title/Abstract] OR "compared"[Title/Abstract] OR "comparing"[Title/Abstract] OR "comparison"[Title/Abstract]))

[Blinding and trial design features]

OR (("double"[Title/Abstract] OR "single"[Title/Abstract] OR "doubly"[Title/Abstract] OR "singly"[Title/Abstract])

AND ("blind"[Title/Abstract] OR "blinded"[Title/Abstract] OR "blindly"[Title/Abstract]))

OR "open label"[Title/Abstract] OR

"double blind procedure"[Title/Abstract] OR

"parallel group*"[Title/Abstract] OR

("crossover"[Title/Abstract] OR "cross over"[Title/Abstract])

[Assignment and allocation]

OR (("assign"[Title/Abstract] OR "match"[Title/Abstract] OR "matched"[Title/Abstract] OR "allocation"[Title/Abstract])

AND ("alternate"[Title/Abstract] OR "group*"[Title/Abstract] OR "intervention*"[Title/Abstract] OR "patient*"[Title/Abstract] OR "subject*"[Title/Abstract] OR "participant*"[Title/Abstract]))

OR ("assigned"[Title/Abstract] OR "allocated"[Title/Abstract])

OR ("controlled"[Title/Abstract] AND ("study"[Title/Abstract] OR "design"[Title/Abstract] OR "trial"[Title/Abstract]))

[Participants and human experimentation]

OR ("volunteer"[Title/Abstract] OR "volunteers"[Title/Abstract])

OR "human experimentation"[MeSH Terms]

OR "trial"[Title/Abstract]

[Economic evaluation and outcome assessment]

OR ("outcome assessment"[Title/Abstract] OR

"health care cost"[Title/Abstract] OR

"primary health care"[Title/Abstract] OR

"economics"[Title/Abstract] OR

"cost*"[Title/Abstract] OR

"cost analysis"[Title/Abstract] OR

"expenditure"[Title/Abstract] OR

"benefit-to-cost"[Title/Abstract] OR

"cost-benefit"[Title/Abstract] OR

"cost-benefit"[Title/Abstract] OR

"cost-consequence"[Title/Abstract] OR

"cost effective*"[Title/Abstract] OR

"cost effective*"[Title/Abstract] OR

"cost utility"[Title/Abstract] OR

"economic evaluation"[Title/Abstract] OR

("outcome assessment, health care"[MeSH Terms] OR

"economics"[MeSH Terms] OR

"costs and cost analysis"[MeSH Terms] OR

"cost benefit analysis"[MeSH Terms] OR

"cost benefit analysis"[MeSH Terms]))

### **Supplementary file 3: Full list of outcome categories, total number reporting at least one outcome and fraction showing significant difference (moderate-low risk of bias only)**

**Table: Number of studies out of total (n=315) reporting domain/subdomain and fraction of studies with moderate-low risk of bias (n=285) showing significant difference**

| **Domain/ subdomain** | **Number of studies out of total (n=315) reporting at least one outcome (%)** | **Fraction of studies* with moderate-low risk of bias (n=285) showing significant difference (%)** |
| --- | --- | --- |
| ***Combined across multiple levels*** |  |  |
| Social inclusion | 13 (4·1) | 3/7 (42·9) |
| Stigma and discrimination | 13 (4·1) | 4/6 (66·0) |
| Human rights abuses | 6 (1·9) | 0/3 (0·7) |
| ***Individual level*** |  |  |
| 1. Sense of belonging in one's community and social acceptance [social inclusion] | 0 (0·0) | 0/0 |
| 2. Sense of agency and capacity to choose whether to participate [social inclusion] | 0 (0·0) | 0/0 |
| 3. Personal recovery [personal recovery] | 14 (4·4) | 3/6 (50·0) |
| 4. Empowerment [personal recovery] | 1 (0·3) | 1/1 (100·0) |
| 5. Self-stigma [stigma and discrimination] | 9 (2·9) | 3/4 (75·0) |
| 6. Quality of life | 41 (13·0) | 20/26 (76·9) |
| 7. Individual general and social functioning | 118 (37·5) | 53/67 (79·1) |
| a) Functioning (general) | 72 (22·9) | 32/43 (74·4) |
| b) Functioning (social) | 65 (20·6) | 29/34 (85·3) |
| 8. Individual mental health & substance use | 197 (62·5) | 91/100 (91·0) |
| a) Mental health (relapse or hospital admission) | 53 (16·8) | 20/28 (71·4) |
| b) Mental health (global symptom severity) | 76 (24·1) | 31/41 (75·6) |
| c) Mental health (psychosis symptom severity) | 18 (5·7) | 12/14 (85·7) |
| d) Mental health (psychosis negative symptom severity) | 76 (24·1) | 33/40 (82·5) |
| e) Mental health (psychosis positive symptom severity) | 80 (25·4) | 35/42 (83·3) |
| f) Mental health (depression/anxiety/distress symptom severity) | 19 (6·0) | 8/11 (72·7) |
| g) Mental health (suicidality) | 3 (1·0) | 1/2 (50·0) |
| h) Mental health (cognitive function) | 28 (8·9) | 13/14 (92·9) |
| i) Mental health (wellbeing) | 9 (2·9) | 3/3 (100·0) |
| j) Mental health (insight) | 19 (6·0) | 9/10 (90·0) |
| k) Mental health (psychological domains) | 5 (1·6) | 2/2 (100·0) |
| l) Substance use | 1 (0·3) | 0/0 |
| 9. Individual physical health & mortality | 12 (3·8) | 6/10 (60·0) |
| a) Physical health (nutritional) | 1 (0·3) | 0/1 (0·0) |
| b) Physical health (cardiometabolic) | 8 (2·5) | 4/6 (66·7) |
| c) Mortality | 3 (1·0) | 1/3 (33·3) |
| d) Physical health (other) | 1 (0·3) | 1/1 (100·0) |
| 10. Adherence to medication | 62 (19·7) | 21/26 (80·8) |
| 11. Knowledge | 7 (2·2) | 2/3 (66·7) |
| 12. Costs | 2 (0·6) | 0/2 (0·0) |
| ***Carer/family level*** |  |  |
| 1. Inclusion in family life [social inclusion] | 0 (0·0) | 0/0 |
| 2. Stigma from family towards people living with psychosis [stigma and discrimination] | 5 (1·6) | 1/1 (100·0) |
| 3. Human rights abuses by family towards people living with psychosis [human rights abuses] | 3 (1·0) | 0/1 (0·0) |
| 4. Family functioning & communication | 21 (6·7) | 6/8 (75·0) |
| a) Family functioning | 14 (4·4) |  |
| b) Family communication | 5 (1·6) |  |
| c) Family caregiving | 3 (1·0) |  |
| 5. Empowerment of carer/family member(s) | 1 (0·3) | 0/0 |
| 6. Quality of life of carer/family member(s) | 7 (2·2) | 3/3 (100·0) |
| 7. Mental health of carer/family member(s) | 10 (3·2) | 3/8 (37·5) |
| 8. Physical health of carer/family member(s) | 1 (0·3) | 1/1 (100·0) |
| 9. Burden on carer/family member(s) | 33 (10·5) | 8/12 (66·7) |
| 10. Knowledge of carer/family member(s) | 10 (3·2) | 5/5 (100·0) |
| 11. Household costs | 9 (2·9) | 1/3 (33·3) |
| ***Level of services or organisations*** |  |  |
| 1. Involvement of people living with psychosis in services/organisations [social inclusion] | 0 (0·0) | 0/0 |
| 2. Structural stigma towards people living with psychosis [stigma and discrimination] | 0 (0·0) | 0/0 |
| 3. Human rights abuses within services & organisations towards people living with psychosis [human rights abuses] | 3 (1·0) | 0/2 (0·0) |
| 4. Engagement of people living with psychosis in services | 4 (1·3) | 1/3 (33·3) |
| 5. Provider stigma towards people living with psychosis [stigma and discrimination] | 1 (0·3) | 0/0 |
| 6. Access to services for people living with psychosis | 0 (0) | 0 |
| 7. Detection and treatment coverage | 2 (0·6) | 1/1 (100·0) |
| 8. Provider competence | 0 (0·0) | 0/0 |
| 9. Involvement of carer/family member(s) in services/organisations | 0 (0·0) | 0/0 |
| 10. Engagement of carer/family member(s) & satisfaction | 20 (6·3) | 1/8 (12·5) |
| 11. Service or system costs | 11 (3·5) | 2/6 (33·3) |
| ***Community level*** |  |  |
| 1. Belonging & acceptance of people living with psychosis [social inclusion] | 0 (0·0) | 0/0 |
| 2. Participation in social life [social inclusion] | 4 (1·3) | 0/2 (0·0) |
| 3. Participation in political life [social inclusion] | 1 (0·3) | 0/0 |
| 4. Participation in economic life [social inclusion] | 10 (3·2) | 3/5 (60·0) |
| 5. Participation in education [social inclusion] | 1 (0·3) | 1/1 (100·0) |
| 6. Public stigma towards people living with psychosis [stigma and discrimination] | 9 (2·9) | 3/5 (60·0) |
| 7. Human rights abuses towards people living with psychosis [human rights abuses] | 1 (0·3) | 0/1 (0·0) |
| 8. Mobilisation of resources by community for people living with psychosis | 0 (0·0) | 0/0 |
| 9. Social support for people living with psychosis from community members | 5 (1·6) | 3/4 (75·0) |
| 10. Affiliate stigma towards carer/family member(s) | 3 (1·0) | 0/0 |
| 11. Employment of carer/family member(s) | 0 (0·0) | 0/0 |
| 12. Education of carer/family member(s) | 0 (0·0) | 0/0 |
| 13. Social support for carer/family member(s) | 1 (0·3) | 0/0 |

*Excluding pilot/feasibility and head to head designs

### **Supplementary file 4: Pre-piloted google sheet for data extraction**

*Format edited for supplementary material.*

| Title of study |  |
| --- | --- |
| Publication title |  |
| Author (First 3) |  |
| Year Published |  |
| Data extractor |  |
| Country |  |
| Study setting [drop down] | Urban, Rural, Mixed, Not specified |
| Other study setting details |  |
| Intervention setting [drop down] | Hospital inpatient, Hospital outpatient, Primary care, Traditional/faith healers, Community, Other |
| Other intervention setting details |  |
| Study design [drop down] | Individual randomised control trial, Cluster randomised control trial, Non-randomised trial, Before-after study, Interrupted time series, Naturalistic study, Other |
| Study design (if other) |  |
| Who is the intervention targeting? [tick all that apply] | Individual |
|  | Family/caregiver |
| Intervention components [tick all that apply] | Antipsychotic medication |
|  | Herbal Remedy |
|  | Case management |
|  | Community-based rehabilitation (CBR) |
|  | Community intervention |
|  | Individual rehabilitation |
|  | Social skills training |
|  | Life skills training |
|  | Recreational (leisure) intervention |
|  | Befriending |
|  | Individual psychoeducation |
|  | Medication adherence interventions |
|  | Individual psychological intervention |
|  | Cognitive remediation/enhancement |
|  | Avatar Therapy |
|  | Individual peer support |
|  | Family/caregiver - psychoeducation |
|  | Family/caregiver - psychological intervention |
|  | Family/caregiver - other |
|  | Group - peer support |
|  | Group - psychological therapy |
|  | Art/music/dance therapy |
|  | Psychodrama |
|  | Physical health intervention |
|  | Economic intervention e.g. cash transfer |
|  | Livelihood/employment intervention |
|  | Empowerment intervention |
|  | Anti-stigma intervention |
|  | mhGAP/Integrated PHC |
|  | Outpatient mental health care |
|  | Service Intervention |
|  | OTHER intervention not listed |
| Who is delivering the intervention? [tick all that apply] | Psychiatrist |
|  | Psychologist |
|  | Social worker |
|  | Occupational therapist |
|  | Psychiatric nurse |
|  | Doctor |
|  | Nurse |
|  | Clinical/health officer |
|  | Community health worker |
|  | Peer |
|  | Lay-worker (non-health) |
|  | Other |
| How much training did they receive? |  |
| Additional details about intervention |  |
| Control condition (if applicable) |  |
| Recruitment (sampling frame) |  |
| Recruitment (sampling method) |  |
| Informed consent |  |
| Diagnosis |  |
| Eligibility criteria |  |
| Sample size (Intervention group) |  |
| Sample size (Control group) |  |
| Gender ration |  |
| Average age |  |
| Education level |  |
| % substance use disorder |  |
| Assessment time points [tick all that apply] | Baseline |
|  | Midpoint(s) during intervention |
|  | Immediately post-intervention (within 2 weeks) |
|  | Follow up post intervention: less than 12 months |
|  | Follow up post intervention: 12 months or more |
| *Outcomes* | *List measure and state if statistically significant (direction of benefit/harm) or non-statistically significant* |
| Mental health outcomes | Psychosis symptom severity |
|  | Remission |
|  | Relapse |
|  | Hospital admission |
|  | Number of episodes |
|  | Cognition (e.g. memory, attention) |
|  | Depressive symptom severity |
|  | Anxiety symptom severity |
|  | Mental distress |
|  | Wellbeing/mental health |
|  | Suicidality |
|  | Violence/aggressive behaviour |
|  | Other |
| Substance misuse outcomes | Alcohol use |
|  | Other substance use |
| Physical health outcomes | Mortality |
|  | Undernutrition/malnutrition |
|  | Overweight/obesity |
|  | Cardiovascular risks (e.g. BP, cholesterol, waist circumference) |
|  | Non-communicable disease (e.g. cardiovascular disease, respiratory disorders, hypertension, diabetes) |
|  | Communicable disease (e.g. TB, HIV, malaria, other) |
|  | Injury/accidents |
|  | Physical health awareness/knowledge |
|  | Physical health self-management |
|  | Other |
| Functioning outcomes | Capabilities |
|  | Days out of role |
|  | Severity of disability/functional impairment (Includes social functioning, work functioning, activities of daily living) |
|  | Other |
| Employment, livelihood and economic outcomes | Employed |
|  | Time in employment |
|  | Quality of employment/work |
|  | Socioeconomic status |
|  | Food security |
|  | Out of pocket healthcare costs |
|  | Lost time and productivity costs |
|  | Livelihood |
|  | Other |
| Social outcomes | Social skills |
|  | Social networks/relationships |
|  | Social support |
|  | Social capital |
|  | Other |
| Education outcomes | Time in education |
|  | Academic achievement |
|  | Absenteeism |
|  | Drop-out from education |
|  | Other |
| Empowerment, stigma and human rights outcomes | Decision-making about care |
|  | Self-management |
|  | Discrimination |
|  | Stigma/self-stigma |
|  | Violent victimisation |
|  | Coercion/restraint |
|  | Other human rights abuses |
|  | Advocacy |
|  | Other |
| Social inclusion | Access/inclusion to community activities |
|  | Access/inclusion to community resources |
|  | Access to livelihood/economic opportunities |
|  | Access to education |
|  | Access to healthcare |
|  | Other |
| Recovery outcomes | Personal/subjective recovery measure |
|  | Hope measure |
|  | Other |
| Quality of life outcomes | Quality of life scale |
|  | Life satisfaction measure |
|  | Other |
| Engagement/adherence outcomes | Engagement |
|  | Satisfaction |
|  | Treatment adherence |
| Caregiver outcomes | Mental health |
|  | Physical health and mortality |
|  | Social (social functioning, status, networks, support, capital) |
|  | Economic/ livelihood impacts (includes catastrophic costs) for an individual caregiver |
|  | Caregiver burden |
|  | Other |
| Family or household level outcomes | Family communication or interaction (e.g. EE), stigmatising attitudes, abuse |
|  | Family involvement (activities done to reflect family's involvement in the person's recovery and care) |
|  | Household economic/food security at level of the household (including catastrophic costs) |
| System outcomes | Involvement of people with lived experience in service development, improvement, governance |
|  | Equitable access |
|  | Affordability |
|  | Quality includes timeliness, safety, etc |
|  | Acceptability, satisfaction, appropriateness |

### **Supplementary file 5: Studies reporting prioritised outcomes**

**Table 1: Studies in psychiatric/not specified inpatient settings with moderate-low risk of bias reporting prioritised outcomes**

| **Author** | **Year** | **Country** | **Design** | **Baseline total** | **Intervention** | **Platform** | **Priority outcome(s) reported** |
| --- | --- | --- | --- | --- | --- | --- | --- |
| Meng et al^1^ | 2005 | China | Controlled clinical trial | 100 | Group art therapy | Hospital (not-specified) inpatient | Self-stigma (SD) |
| Chen et al^2^ | 2003 | China | Randomised controlled trial | 64 | Social skills training | Psychiatric hospital inpatient | Employment (SD) |
| Tang et al^3^ | 2023 | China | Randomised controlled trial | 54 | Expressive writing | Psychiatric hospital inpatient | Personal recovery (Hope) (SD) |
| Alhadidi et al^4^ | 2023 | Jordan | Controlled clinical trial | 122 | Psychoeducation | Hospital (not-specified) inpatient | Self-stigma (SD) |

SD = statistically significant difference

**Table 2: Studies with high risk of bias reporting prioritised outcomes (any platform)**

| **Author** | **Year** |  | **Country** | **Design** | **Baseline total** | **Intervention** | **Platform** | **Priority outcome(s) reported** |
| --- | --- | --- | --- | --- | --- | --- | --- | --- |
| Sitthimongkol et al^5^ | 2007 | 19 | Thailand | Uncontrolled before-after study | 18 | Psychoeducation and mutual support for family members | Hospital (not-specified) out-patient | Family stigma (NS) |
| Lu et al^6^ | 2014 | 35 | China | Controlled clinical trial | 60 | Inpatient nursing care intervention | Psychiatric hospital in-patient | Self-stigma (SD) |
| Zhou et al^7^ | 2002 | 48 | China | Controlled clinical trial | 24 | Psychodrama therapy | Psychiatric hospital in-patient | Personal recovery (self-esteem) (SD) |
| Tang et al^8^ | 2006 | 49 | China | Randomised controlled trial (individual) | 100 | Psychoeducation for family and people living with psychosis | Psychiatric hospital in-patient | Personal recovery (SD) |
| Hanlon et al^9^ | 2020 | 57 | Ethiopia | Uncontrolled before-after study | 300 | Integrated mental health care | Primary care | Public stigma (SD) |
| Gutierrez-Maldonado et al^10^ | 2009 | 67 | Chile | Controlled clinical trial | 41 | Multi-family psychoeducation | Psychiatric hospital out-patient | Family stigma (SD) |
| Suryani et al^11^ | 2011 | 71 | Indonesia | Uncontrolled before-after study | 23 | Community treatment combining depot medication with a psycho-spiritual approach | Mixed | Human rights abuses (restraint) (SD) |
| Bachtiar et al^12^ | 2019 | 112 | Indonesia | Two group pre-post | 82 | Multi-group education and training intervention | Community | Self-stigma (SD), Family stigma (SD), Public stigma (SD) |
| Guan et al^13^ | 2015 | 131 | China | Uncontrolled before-after study | 266 | Unlocking people living with psychosis in restraints at home and provision of community care | Community | Productive labour/work (SD) |
| Chatterjee et al^14^ | 2009 | 164 | India | Uncontrolled before-after study | 256 | Community-based rehabilitation and psychiatric outreach | Community | Participation in economic, social and political life (SD) |
| Yulina et al^15^ | 2020 | 169 | Indonesia | Two group pre-post | 54 | Group-based mindfulness therapy | Psychiatric hospital in-patient | Personal recovery (SD) |
| Zavradashvili et al^16^ | 2010 | 176 | Georgia | Uncontrolled before-after study | 26 | Assertive community treatment | Community | Participation in economic life (SD) |
| Worakul et al^17^ | 2007 | 187 | Thailand | Uncontrolled before-after study | 91 | Psycho-education for caregivers | Psychiatric hospital out-patient | Family stigma (SD) |
| Cao et al^18^ | 2015 | 200 | China | Controlled clinical trial | 65 | Cognitive therapy | Psychiatric hospital out-patient | Self-stigma (SD) |
| Li et al^19^ | 2015 | 203 | China | Controlled clinical trial | 95 | Cognitive behavioural therapy | Psychiatric hospital out-patient | Self-stigma (SD) |
| Liu et al^20^ | 2013 | 204 | China | Randomised controlled trial (individual) | 60 | ‘Hope-based’ intervention | Psychiatric hospital out-patient | Personal recovery (SD), Self-stigma (SD) |
| Lu et al^21^ | 2004 | 205 | China | Controlled clinical trial | 112 | Social skills training | Psychiatric hospital in-patient | Personal recovery (SD) |
| Ponnuchamy et al^22^ | 2011 | 211 | India | Uncontrolled before-after study | 60 | Structured social work | Community | Participation in social life (SD) |
| Liu et al^23^ | 2004 | 257 | China | Controlled clinical trial | 118 | Family intervention | Mixed | Personal recovery (SD) |
| Ngoc et al^24^ | 2016 | 291 | Vietnam | Controlled clinical trial | 59 | Psychoeducation for family and people living with psychosis | Psychiatric hospital in-patient | Public stigma (SD) |
| Bhawana et al^25^ | 2022 | 310 | India | Uncontrolled before-after study | 120 | Psychoeducation for family and people living with psychosis | Psychiatric hospital in-patient | Personal recovery (SD) |

NS = non-statistically significant difference; SD = statistically significant difference indicating potential benefit

### **Supplementary file 6: Studies reporting outcomes across multiple levels of the socio-ecological model**

**Table 1: Studies in psychiatric/not specified inpatient settings with moderate-low risk of bias reporting outcomes across multiple levels of the socio-ecological model**

| **Author** | **Year** | **Country** | **Design** | **Baseline total** | **Intervention** | **Platform** | **Individual** | **Family** | **Organisations** | **Community** |
| --- | --- | --- | --- | --- | --- | --- | --- | --- | --- | --- |
| Shen et al^26^ | 2013 | China | Randomised controlled trial (individual) | 101 | Rehabilitation intervention | Hospital (not-specified) in-patient | Functioning (SD), Quality of life (SD) | Family functioning (SD) |  |  |
| Chen et al^27^ | 2003 | China | Randomised controlled trial (individual) | 64 | Social-skills training | Psychiatric hospital in-patient | Functioning (SD), Mental health (SD) |  |  | Participation in economic life (SD) |
| Ma et al^28^ | 2004 | China | Randomised controlled trial (individual) | 221 | Family intervention | Psychiatric hospital in-patient | Functioning (SD), Mental health (SD) | Family functioning (SD) |  |  |
| Tang et al^3^ | 2023 | China | Randomised controlled trial (individual) | 54 | Expressive Writing | Psychiatric hospital in-patient | Personal recovery (SD), Quality of life (SD) |  |  | Public stigma towards people living with psychosis (SD) |

SD = statistically significant difference indicating potential benefit

**Table 2: Studies with high risk of bias reporting outcomes across multiple levels of the socio-ecological model (any platform)**

| **Author** | **Year** | **Country** | **Design** | **Baseline total** | **Intervention** | **Platform** | **Individual** | **Family** | **Organisations** | **Community** |
| --- | --- | --- | --- | --- | --- | --- | --- | --- | --- | --- |
| Amaresha et al^29^ | 2017 | India | Controlled clinical trial | 80 | Psychoeducation | Psychiatric hospital in-patient and out-patient |  | Burden on carer/family member(s) (NS), Family knowledge (SD) |  | Affiliate stigma (SD) |
| Stanley et al^30^ | 2006 | India | Uncontrolled before-after study | 60 | Psychiatric intervention combined with traditional rituals | Social care/community residential | Mental health (SD) | Burden on carer/family member(s) (SD), Quality of life of carer/family member(s) (SD) |  |  |
| Sungur et al^31^ | 2003 | Turkey | Controlled clinical trial | 100 | Case management | Psychiatric hospital out-patient | Mental health (SD) | Burden on carer/family member(s) (SD) |  |  |
| Sitthimongkol et al^5^ | 2007 | Thailand | Uncontrolled before-after study | 18 | Psychoeducation and mutual support for family members | Hospital (not-specified) out-patient | Functioning (NS/SD) | Empowerment of carer/family member(s) (NS), Family functioning (NS), Stigma from family towards people living with psychosis (SD) |  |  |
| Jiang et al^32^ | 2014 | China | Randomised controlled trial (individual) | 143 | Skills training | Community | Functioning (SD), Mental health (SD) | Family functioning (SD) |  |  |
| Hanlon et al^9^ | 2020 | Ethiopia | Uncontrolled before-after study | 300 | Integrated mental health care (PRIME) | Primary health care/community health centre | Functioning (SD), Mental health (SD), Substance use (SD) | Human rights abuses by family towards people living with psychosis (SD) |  | Public stigma towards people living with psychosis (SD) |
| Paranthaman et al^33^ | 2010 | Malaysia | Two group pre-post | 218 | Group psychoeducation for caregivers | Community mental health service | Mental health (NS) | Burden on carer/family member(s) (NS), Knowledge of carer/family member(s) (SD) |  |  |
| Nakku et al^34^ | 2019 | Uganda | Uncontrolled before-after study | 51 | Integrated mental health care (PRIME) | Primary health care/community health centre | Functioning (SD) | Household costs (SD) | Service or system costs (SD) |  |
| Malla et al^35^ | 2020 | India | Uncontrolled before-after study | 333 | Early intervention services | Psychiatric hospital out-patient | Mental health (SD) | Family caregiving (SD) |  |  |
| Sadath et al^36^ | 2017 | India | Controlled clinical trial | 71 | Psychoeducation and psychosocial intervention | Psychiatric hospital out-patient |  | Family communication (NS) |  | Social support for carer/family member(s) (NS) |
| Jordans et al^37^ | 2019 | Nepal | Uncontrolled before-after study | 95 | Integrated mental health care (PRIME) | Primary health care/community health centre |  | Household costs (NS) | Detection and treatment coverage (SD), Service or system costs (NS) |  |
| Martínez et al^38^ | 2005 | Mexico | Uncontrolled before-after study | 78 | Rehabilitation programme | Psychiatric hospital out-patient | Functioning (SD), Mental health (NS) | Family communication (SD) |  |  |
| Özdemir et al^39^ | 2017 | Turkey | Controlled clinical trial | 150 | Psychosocial rehabilitation intervention | Psychiatric hospital out-patient | Adherence (SD), Functioning (SD), Mental health (SD) | Burden on carer/family member(s) (SD) |  |  |
| Bachtiar et al^12^ | 2019 | Indonesia | Two group pre-post | 82 | Multi-group education and training intervention | Community | Self-stigma (SD) | Stigma from family towards people living with psychosis (SD) | Provider stigma towards people living with psychosis (SD) | Public stigma towards people living with psychosis (SD) |
| Doğan et al^40^ | 2004 | Turkey | Uncontrolled before-after study | 23 | Home visits with psychoeducation and skills training for caregivers | Community | Functioning (SD), Quality of life (SD) | Family functioning (SD) |  | Social support for people living with psychosis from community members (SD) |
| Guan et al^13^ | 2015 | China | Uncontrolled before-after study | 266 | "Unlocking" programme | Community | Adherence (SD), Functioning (SD), Mental health (SD) | Burden on carer/family member(s) (SD), Household costs (SD) |  | Participation in economic life (SD), Public stigma towards people living with psychosis (SD) |
| Chatterjee et al^14^ | 2009 | India | Uncontrolled before-after study | 256 | Community-Based Rehabilitation | Community | Functioning (SD) |  |  | Participation in economic life (SD), Participation in political life (SD), Participation in social life (SD) |
| Zavradashvili et al^16^ | 2010 | Georgia | Uncontrolled before-after study | 26 | Assertive community treatment | Community | Adherence (NS), Mental health (SD) |  |  | Participation in economic life (SD) |
| Zhang et al^41^ | 2018 | China | Uncontrolled before-after study | 107 | The un-locking programme | Mixed hospital/community | Functioning (NS), Mental health (NS) | Burden on carer/family member(s) (SD) |  |  |
| Chan et al^42^ | 2007 | China | Controlled clinical trial | 60 | Psychoeducation (people living with psychosis) | Hospital (not-specified) in-patient | Quality of life (NS) | Family functioning (NS) |  |  |
| Zhang et al^43^ | 2008 | China | Controlled clinical trial | 157 | Psychoeducation (people living with psychosis) | Not specified | Adherence (NS), Mental health (SD) | Burden on carer/family member(s) (SD) |  |  |
| Devaramane et al^44^ | 2011 | India | Uncontrolled before-after study | 40 | Psychoeducation (family) | General hospital setting (not-specified) | Mental health (SD) | Burden on carer/family member(s) (SD), Family communication (SD), Family functioning (SD) |  |  |
| Sharma et al^45^ | 2021 | India | Controlled clinical trial | 40 | Psychoeducation (family) | Not specified | Mental health (SD) | Mental health of carer/family member(s) (SD) |  |  |
| Ngoc et al^24^ | 2016 | Vietnam | Controlled clinical trial | 59 | Psychoeducation (people living with psychosis and family) | Psychiatric hospital in-patient | Adherence (SD), Quality of life (NS/SD) |  |  | Affiliate stigma towards carer/family member(s) (SD), Public stigma towards people living with psychosis (SD) |
| Kumar et al^46^ | 2020 | India | Randomised controlled trial (individual) | 66 | Brief Psychosocial Intervention (family/caregiver) | Psychiatric hospital out-patient | Mental health (NS), Quality of life (SD) | Burden on carer/family member(s) (SD), Quality of life of carer/family member(s) (SD) |  |  |
| Sengupta et al^47^ | 2021 | India | Two group pre-post | 10 | Functional Analytic Psychotherapy | Psychiatric hospital in-patient | Mental health (SD) | Mental health (SD) |  |  |
| Bhawana et al^25^ | 2022 | India | Uncontrolled before-after study | 120 | Psychoeducation (people living with psychosis and family) | Psychiatric hospital in-patient | Adherence (SD), Personal recovery (SD) | Burden on carer/family member(s) (SD) |  |  |
| Rahmatnejad et al^48^ | 2022 | Iran | Randomised controlled trial (individual) | 60 | Self-Care Training Based On Acceptance and Commitment Therapy designed for caregivers | Not specified |  | Adherence (SD), Personal recovery (SD) |  | Affiliate stigma (SD) |
| Liu et al^49^ | 2022 | China | Uncontrolled before-after study | 100 | Injectable antipsychotic intervention | Psychiatric hospital out-patient |  | Household costs (SD) | Service or system costs (NS) |  |
| Chisholm et al^50^ | 2020 | India | Uncontrolled before-after study | 39 | Integrated mental health care (PRIME) | Primary health care/community health centre |  | Household costs (SD) | Service or system costs (NS) |  |

NS = non-statistically significant difference; SD = statistically significant difference indicating potential benefit

19.	Li; Y.L. Xiong; A.L HLF. Effects of behaviour treatment for schizophrenia patients during recovery stage. 2015.

20.	Liu; G.L WWX, Ma; X. The study of using hoping theory in clinical rehabilitation nursing for patients with schizophrenia. 2013.

21.	Lu; SC LL, Fu; FZ. Effect of social skill training on social functions and happiness degree of patients with schizophrenia. 2004.

22.	Ponnuchamy L. Social Work Intervention for Disability Management of Persons with schizophrenia in India with reference to Rural areas. *The International Journal of Psychosocial Rehabilitation* 2012.

28.	Ma; X LL, Q; H; Liu. Effect of individual system family intervention on the family and social functions in patients with schizophrenia: A randomized control study. 2004.
